# Using Satellite Images and Deep Learning to Identify Associations Between County-Level Mortality and Residential Neighborhood Features Proximal to Schools: A Cross-Sectional Study

**DOI:** 10.1101/2020.10.12.20211755

**Authors:** Joshua J. Levy, Rebecca M. Lebeaux, Anne G. Hoen, Brock C. Christensen, Louis J. Vaickus, Todd A. MacKenzie

## Abstract

**What is the relationship between mortality and satellite images as elucidated through the use of Convolutional Neural Networks?**

**Background:** Following a century of increase, life expectancy in the United States has stagnated and begun to decline in recent decades. Using satellite images and street view images, prior work has demonstrated associations of the built environment with income, education, access to care and health factors such as obesity. However, assessment of learned image feature relationships with variation in crude mortality rate across the United States has been lacking.

**Objective:** We sought to investigate if county-level mortality rates in the U.S. could be predicted from satellite images.

**Methods:** Satellite images of neighborhoods surrounding schools were extracted with the Google Static Maps application programming interface for 430 counties representing approximately 68.9% of the US population. A convolutional neural network was trained using crude mortality rates for each county in 2015 to predict mortality. Learned image features were interpreted using Shapley Additive Feature Explanations, clustered, and compared to mortality and its associated covariate predictors.

**Results:** Predicted mortality from satellite images in a held-out test set of counties was strongly correlated to the true crude mortality rate (Pearson r=0.72). Direct prediction of mortality using a deep learning model across a cross-section of 430 U.S. counties identified key features in the environment (e.g. sidewalks, driveways and hiking trails) associated with lower mortality. Learned image features were clustered, and we identified 10 clusters that were associated with education, income, geographical region, race and age.

**Conclusions:** The application of deep learning techniques to remotely-sensed features of the built environment can serve as a useful predictor of mortality in the United States. Although we identified features that were largely associated with demographic information, future modeling approaches that directly identify image features associated with health-related outcomes have the potential to inform targeted public health interventions.

## Introduction

Life expectancy in the United States has increased dramatically over the past century from 48 years in 1900 to 80 years in 2019. However, the United States has experienced a drop in longevity over the past decade and now ranks 43^rd^ in the world [1–5]. Within the United States, crude mortality rates vary by more than 40%. Factors observed to be associated with mortality rates within the United States include disparities in socioeconomic status [6, 7] and health insurance coverage, [8] as well as obesity, smoking [9, 10] and drug use/opioid abuse [11].

Prior studies have attempted to infer characteristics of the underlying communities by characterizing land use using satellite imagery [12–14]. Recent research has leveraged deep learning approaches to link the built environment to obesity [15, 16], socioeconomic status [17], poverty [18, 19], and other demographic factors [20, 21] by integrating lower level image features into higher order abstractions to make predictions [22, 23]. In addition, the potential for deep learning-based image analysis to characterize a broad range of health exposures was recently reviewed [24]. However, there is limited work that examines the relationship of mortality with the built environment in the United States at a large scale. The identification of information in satellite images associated with mortality predictions could potentially unlock previously unknown, yet related geographic and structural community characteristics that may be used to predict future mortality rates when demographic information is unavailable or possibly inform optimal county/state public health intervention strategies.

We hypothesized that county-scale level mortality rates could be predicted from satellite images. The goals of this study were to determine whether the analysis of satellite images by Convolutional Neural Networks (CNN) could be used to predict county-level mortality rates and uncover salient satellite image features associated with mortality. We also sought to determine if image features are related with county-level measures of income, education, age, sex, race and ethnicity. The main objective of this study was to highlight a proof-of-concept deep learning application that presents strong baseline performance, enabling future work that can evaluate the potential for applying learnt features to the design of public health interventions in the built environment.

## Methods

### Overview

Here, we will provide a brief overview of methods used to train, validate, test, interpret and compare the selected machine learning models for the task of mortality prediction:

1. **Data collection:** We collected death certificate and county-level covariate/demographic information from private access census databases correspondent to 430 US counties, partitioning counties into training, validation and test sets. We downloaded 196 Google Maps satellite images per county.
2. **Predictive Modeling:** We trained, validated and tested three separate models for comparison:
  a. A deep learning model which operates on images to predict the county-level mortality rate for each image. The predicted mortality rates for the images within each county are averaged with a trimmed mean to yield the final county-level mortality predictions.
  b. A linear regression model which can predict county-level mortality from county-level covariate information.
  c. A hybrid deep learning approach which can combine county-level covariates with image features to predict image-specific mortality rates, averaged (trimmed mean) across the county to yield county-level mortality rates.
  d. **Data Scaling Sensitivity Tests** to identify the amount of imaging information needed for optimal predictive accuracy for the image-only deep learning model.
3. **Interpretation Techniques** to identify important and potentially correspondent image and demographic predictors of mortality, using:
  a. **SHAP** to generate a heatmap over each of the images to locate mortality associated image features.
  b. **Standardized regression coefficients and SHAP** to identify the most important demographic predictors of mortality from the regression model.
  c. **Unsupervised dimensionality reduction** to identify correspondence between the demographics and imaging predictors.

Graphical overviews of the aforementioned approaches may be found in Figures 1-2 and in the Supplementary Material, section “Supplementary Overview of All Conducted Analyses”.

**Figure 1:**
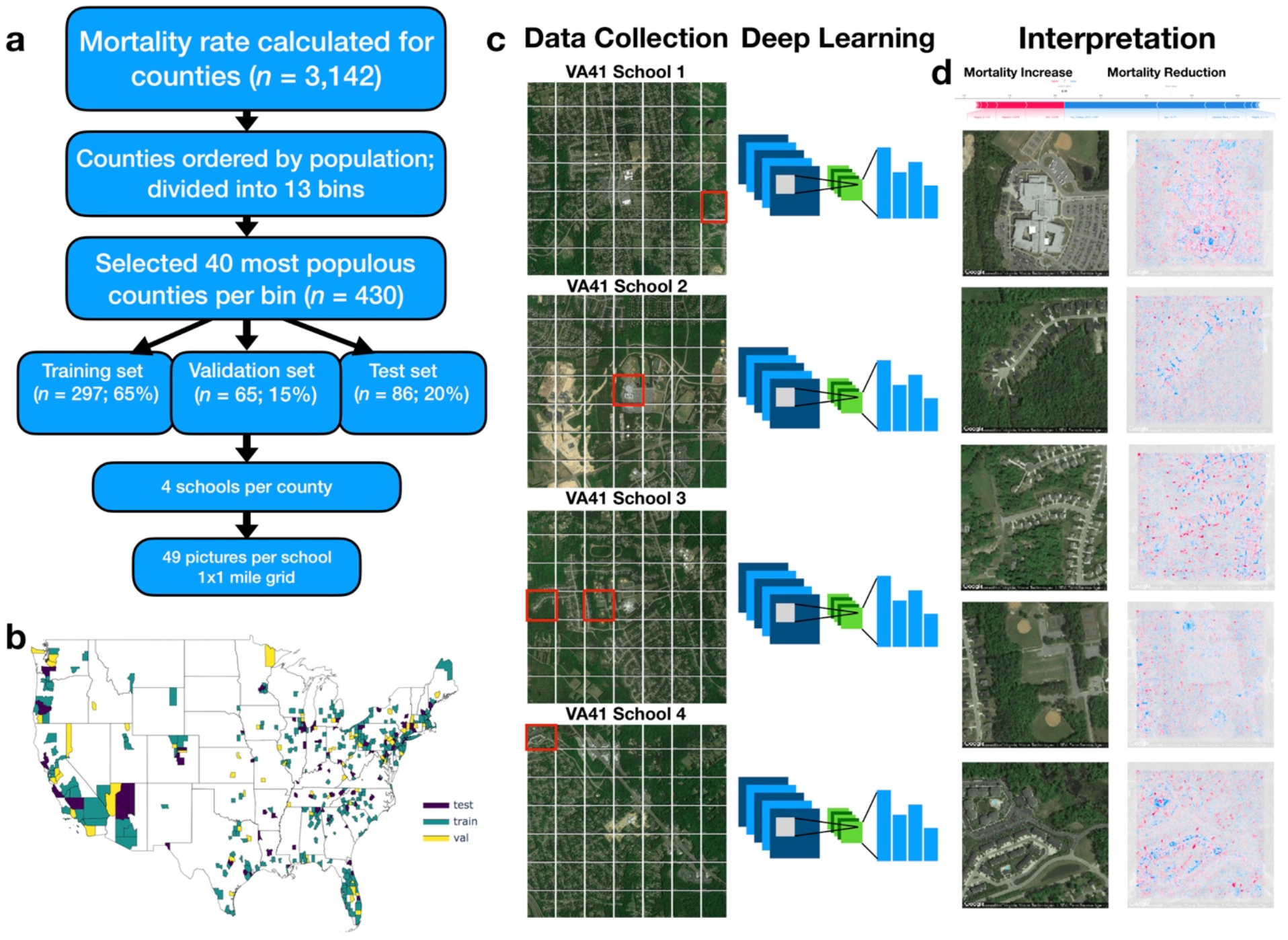
Method Overview: a) Flow diagram depicting selection of counties and sampling scheme for images; 13 mortality bins created from 1000 most populous counties to capture diversity of mortality rates; b) Map of the United States colored by selection of training, testing and validation counties; c) 4 sets of 1×1 mile grid of images (7 images by 7 images) were selected per each county, all images randomized and used to train deep learning model; visual depiction of deep learning: filters slide across each image and pick up on key image features, pooling layers reduce spatial dimensions of image, and output layers of neural network output one mortality rate prediction per image, *ŷ*i; histogram visualization to the right of the neural network represents the set of mortality rate predictions {*ŷ*i | *i*∈{1,2,…,195,196}}corresponding to 196 images per county (49 images per school * 4 schools per county), which are averaged across using a trimmed mean to yield final mortality rates 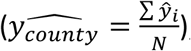; d) SHAP is applied to neural network model and image to generate a heatmap of important image features across each image (red=positively associated with mortality; blue=negatively associated with mortality); these visualizations are compared to important covariates from linear model for each test county (visualized as force plot placed above the heatmap images; where presence of colors red/blue indicates positive/negative associations with mortality).

**Figure 2:**
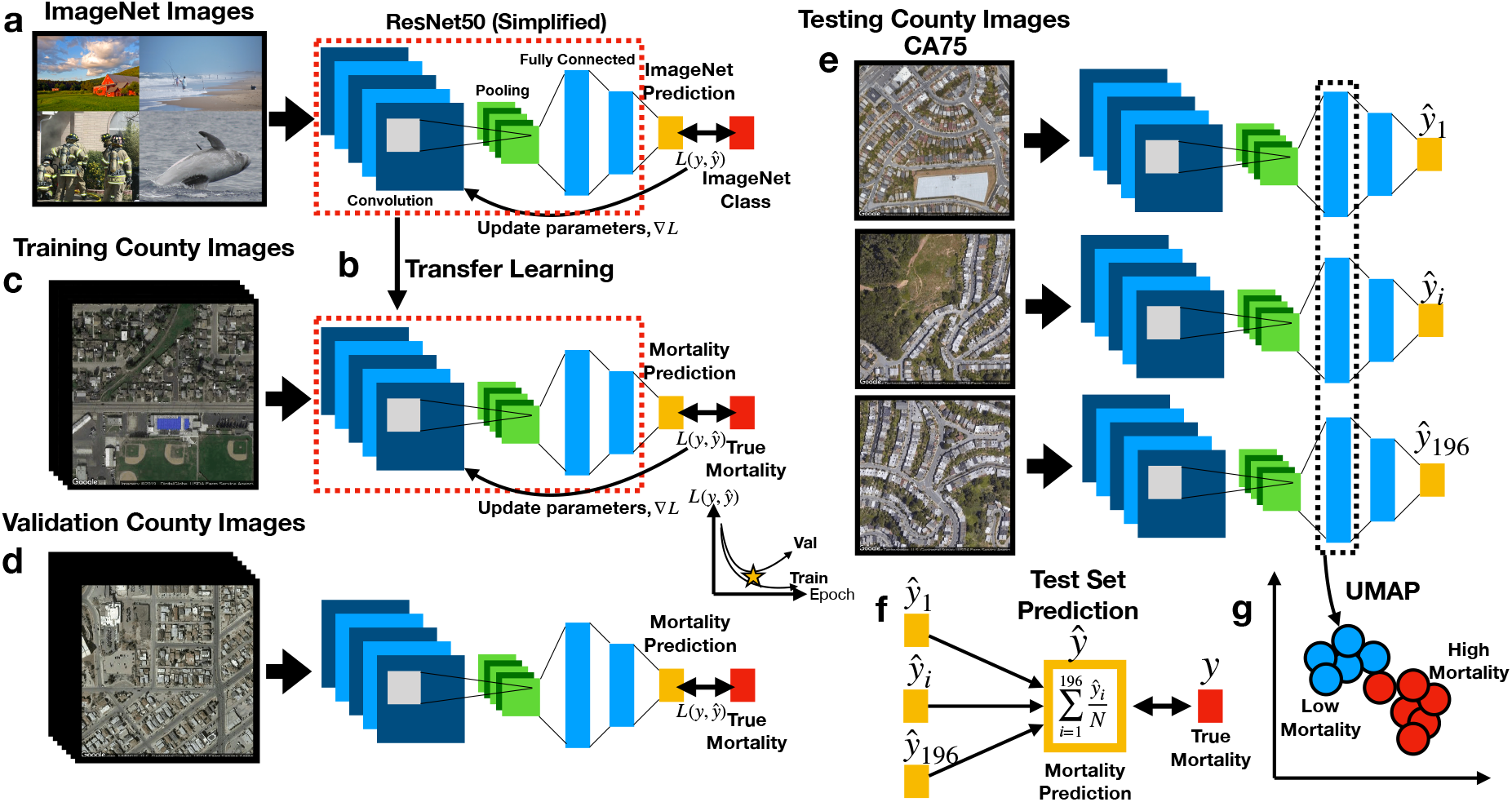
Deep Learning Workflow: a) ResNet50 model is first pre-trained on ImageNet images for the task of distinguishing 1000 different everyday objects (e.g., farm, beach, firemen, whale); ResNet50 is comprised of multiple convolution and pooling layers (only one set shown for simplicity), which extract features and reduce the spatial dimensions of the image respectively, and fully connected layers which represents objects at multiple levels of abstraction using vectors; prediction is compared to ground truth to estimate loss *L*; the gradient is used to update the model parameters; b) transfer learning, where parameters from pretrained model (red dashed box) serve as the initial parameters for training the country-level mortality prediction model; output layer has been replaced with a layer to predict mortality; c) training images pass through ResNet50 to predict county-level mortality, compared to true mortality to update model parameters; d) during training, ResNet50 model predicts mortality from images from validation counties to estimate validation loss, which is compared to training loss over training epochs to determine when training should stop to avoid overfitting; e) trained ResNet50 model predicts county-level mortality on 196 individual images (one mortality rate prediction per image) from each test county; f) for each test county, 196 image-level mortality rate predictions are averaged using a trimmed mean to infer the overall county-level mortality rate; the overall county-level mortality rate is correlated to true county-level mortality rate; g) image features from images of the test counties are extracted from ResNet50 fully connected layers in the form of feature vectors, which are used to generate UMAP plots to demonstrate how mortality and other corresponding covariates (extracted from the demographic data) separate/cluster based on the image features; Spectral clustering is used to assign image features to blue and red clusters, representing low and high mortality related images respectively.

### Data Collection

#### Extraction of County-Level Covariates

We used publicly available county-level mortality data. We used the CDC Wonder database to collect all available death data from 2015 for residents of the 50 United States and the District of Columbia, and matched death data with 2015 county Census population data from the Bureau of Economic Analysis, USDA ERS databases [25–27], and the Surveillance, Epidemiology, and Ends Results (SEER) Program [28]. Crude mortality rates were calculated as the number of deaths in the county divided by the population in the county. Additional county level covariate information from 2015 was collected on age, sex, Hispanic status, race, income, education, and region (Supplementary Table 1).

#### Selection Criteria for Training, Validation and Test Sets

We selected 430 counties from among 3,142 total United States counties in 2015 for inclusion in our study. This subset of representative counties was selected to decrease the computational burden and data storage resources. To reduce the variance of the mortality estimate and to limit potential bias from locale (e.g. rural, urban), we selected the top 1000 most populous counties (restricting to more urban environments) and split these counties into 13 bins containing similar numbers of counties rank ordered by mortality. We selected up to forty of the most populous counties in each bin. These selection criteria were developed to capture a greater proportion of total deaths and represent wider variation in mortality rates, ultimately resulting in the selection of the aforementioned 430 counties (Supplementary Table 2; in-depth description in Supplementary Figure 1). Of the selected counties, 279 counties were randomly placed into the training set (65%), 65 in the validation set (15%), and 86 in the test set (20%). The training set was used to update the parameters of the trained model while the validation set was used to limit the model from overfitting to the training data (Figure 1a-b; Figure 2a-d). The held-out test set represents an application of the modeling approach to unseen counties that had no role in the training of the model.

#### Acquisition of Imaging Data

Satellite imagery data was collected using the Google Static Maps application programming interface (API) to build our deep learning pipeline, similar to Maharana and Nsoesie [15]. First, four schools from each county were randomly selected to serve as points of interest (POI) to sample nearby images. We used schools because they are typically placed in densely populated regions of the county (potentially serving as a representative proxy for residential neighborhoods of each county) and to reduce the computing space usage and required resources at our local computing cluster and acknowledge that the selection of schools as a POI presents a limited view of the entire county. We obtained geographic coordinates for the schools in our study from the National Center for Education Statistics [29], and divided the 1 square mile area surrounding each school into an evenly spaced 7 images by 7 images grid (extracting 196 total images per county) (Figure 1c). The zoom level for each image was set to 17, and image dimensions were set to 400 pixels by 400 pixels to provide enough detail to make out street patterns and cover the space between the selected images of the grid. 84,280 satellite images were downloaded using the Google Static Maps API. Images were downloaded between July and September 2019, representing a collection of images acquired April 2018 to December 2018.

This study was exempt from institutional review board approval as we accessed previously collected data and could not identify individuals. Our study followed the Strengthening the Reporting of Observational Studies in Epidemiology (STROBE) guidelines for cross-sectional studies where appropriate [30].

### Predictive Modeling

#### Deep Learning Model Training

Deep neural networks have been used in a wide range of health-related applications [31, 32]. Convolutional Neural Networks (CNN) slide filters across images to pick up on low level features such as edges or curves and then expand the visual field to pick up higher-order constructs (Figure 1c) [33]. The particular variant of CNN that was used to predict mortality on the satellite images was ResNet-50 [34], pretrained on the ImageNet database to recognize over 1000 different objects [35], some of which may have features corresponding with our satellite image set (Figure 2a-b). We performed transfer learning, which applies knowledge gained from ImageNet to initialize the parameters of our model. These model parameters were updated to minimize the divergence between the true and predicted mortality rates using the negative Poisson log likelihood; training the model with this objective (models the outcome as a rate) allowed us to use each image to directly predict the mortality of its respective county. Training images were randomly cropped, rotated, resized, and flipped to improve the generalizability of the approach (Figure 2c). Predicted mortality rates from the individual images were averaged across the images for each county using a trimmed mean to comprise the final county-level prediction. A validation set of images from the validation counties were used to select optimal hyperparameters (e.g., learning rate, weight decay, early stopping criterion) to avoid memorization of the training data (Figure 2d). After a coarse hyperparameter search, the validation set was used to terminate the learning process at 5 training iterations and identify the ideal learning rate for the model, 1e-4. After training our model on images from 279 counties, we evaluated the model on images from the remaining test counties and averaged the predicted image-level mortality rates across each county using a trimmed mean to derive county mortality estimates (Figure 2e-f). The deep learning models developed using Python 3.6, utilized the PyTorch 1.3.0 framework and were trained using K80 Graphics Processing Units (GPUs).

#### Linear Regression on County-Level Covariates

A linear regression model to predict mortality using county level demographic characteristics was fit to data from both the training and validation sets, weighted by county population size (to reduce the variance of the parameter estimates), and evaluated on the test set as a comparison method versus the satellite image approach (Supplementary Figure 2b). We note that the goal of this study was to provide a benchmark for how well the CNN could predict mortality and to use covariates to help contextualize what the deep learning model is “seeing”.

#### Hybrid Image-Covariate Deep Learning Approach

We also combined demographic information with satellite imagery data to test whether adjustment for covariates can improve prediction of mortality from satellite images (Supplementary Figure 2c, see Supplementary Materials section “Covariate Adjustment During Deep Learning Model Training and Evaluation”).

#### Data Scaling Sensitivity Tests

A sensitivity analysis was utilized to decide the ideal number of schools and sampling area around the schools for the deep learning model (Supplementary Figure 2e-f). Preliminary tests from our modeling approach indicated that mortality prediction performance increases with the number of schools and area around the school sampled. However, performance saturates as the number of images in each county approaches 196, thereby warranting selection of images contained within a one-mile square area around schools to maximize the potential utility of assessing schools as a POI and limit the amount of noise created beyond surveying neighborhoods in the immediate vicinity of schools. See Supplementary Materials, section “Effects of Sampling Larger Residential Areas Around Schools and Dataset Size on Predictions”.

### Model Interpretation

#### Image Interpretations with SHAP

Shapley additive feature explanations (SHAP) [36] is an analytical technique that explains complex models using a simpler surrogate model for each testing instance. We applied SHAP to the images of the test counties to form pixel-wise associations with increases or reductions in mortality, hotspots in these images denote important mortality-associated objects that the model has learned (Figure 1d). We also convolved learned CNN image filters over select images to further demonstrate which features of the built environment were utilized by deep learning model.

#### Important County-Level Covariates with SHAP and Standardized Regression Coefficients

SHAP was applied to identify county-specific important covariate predictors (Figure 1d) from the linear model, similar to how the SHAP image approach could identify important image-specific predictors. Overall effect estimates were reported using unstandardized regression coefficients. Standardized regression coefficients from the linear regression model served to explain the overall top mortality predictors.

#### Unsupervised dimensionality reduction to identify correspondent demographic and image predictors

Correspondence between the covariate mortality predictors for each county and the image information was assessed through *embedding* of deep learning image features. Neural networks compress high-dimensional image data into lower dimensional representations in the process of making a prediction (Figure 2g, Supplementary Figure 2d). The output of an intermediate layer of the network was extracted from each image to form *embeddings* – reduced dimensional representations of the data in the form of vectors. These *embeddings* (represented by 1000-dimensional vectors) could demonstrate how overall features of the images cluster and correspond with mortality or other demographics. We applied UMAP [37], an unsupervised dimensionality reduction technique, to visualize extract image features and identify clustering by images using scatterplots (one image is a point in the scatterplot). We overlaid the actual images themselves for each of the points, then true mortality and other covariates such as education and aging to visually demonstrate separation/clustering of important image-associated demographic and mortality characteristics as learned by the deep learning model. To examine specific associations between the pertinent image features (i.e. embeddings), we clustered image features using the Spectral Clustering approach [38] and found 10 clusters of images. We averaged the covariate information associated with each of the images across each grouping to yield characteristic covariate descriptors for each cluster. Weighted pairwise t-tests on mean differences between select covariates between all pairs of clusters demonstrated associations between extracted image features with covariate mortality predictors. As a direct means for assessing the relationship between deep learning predicted mortality and the covariates, we also regressed the image predicted mortality against each of these covariates.

## Results

The training, validation and test counties included 1,721,052 deaths per 217,938,597 individuals in the 2015 U.S. population, a crude mortality rate of 7.90 deaths per 1,000 U.S. residents (Supplementary Table 1).

### Predictive Modeling Results

Our deep learning model was able to accurately predict mortality, with a Pearson correlation coefficient of 0.72 (R^2^=51%) between the predicted and true mortality (Figure 3a) on a held-out test set. The linear regression model using county demographics also accurately predicted mortality; the R^2^ was 90% (0.95 correlation) between covariates and the mortality rate (Figure 3b). Our covariate adjusted deep learning model appeared to improve mortality predictive performance (Test Set Pearson r=0.83, *P*<0.001; Validation Set Pearson r=0.87, *P*<0.001) but was unable to surpass the performance achieved using only the demographic characteristics (Supplementary Materials, section “Covariate Adjustment During Deep Learning Model Training and Evaluation”) Sensitivity analyses over the included number of schools and area around schools indicated that mortality predictive performance tends to increase with the number of schools and area sampled around each school. However, predictive performance appeared to level-off after selection of 75 images per county (3 schools per county; 25 images per school; each image occupies 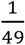 square mile) (Supplementary Materials, section “Effects of Sampling Larger Residential Areas Around Schools and Dataset Size on Predictions”).

**Figure 3:**
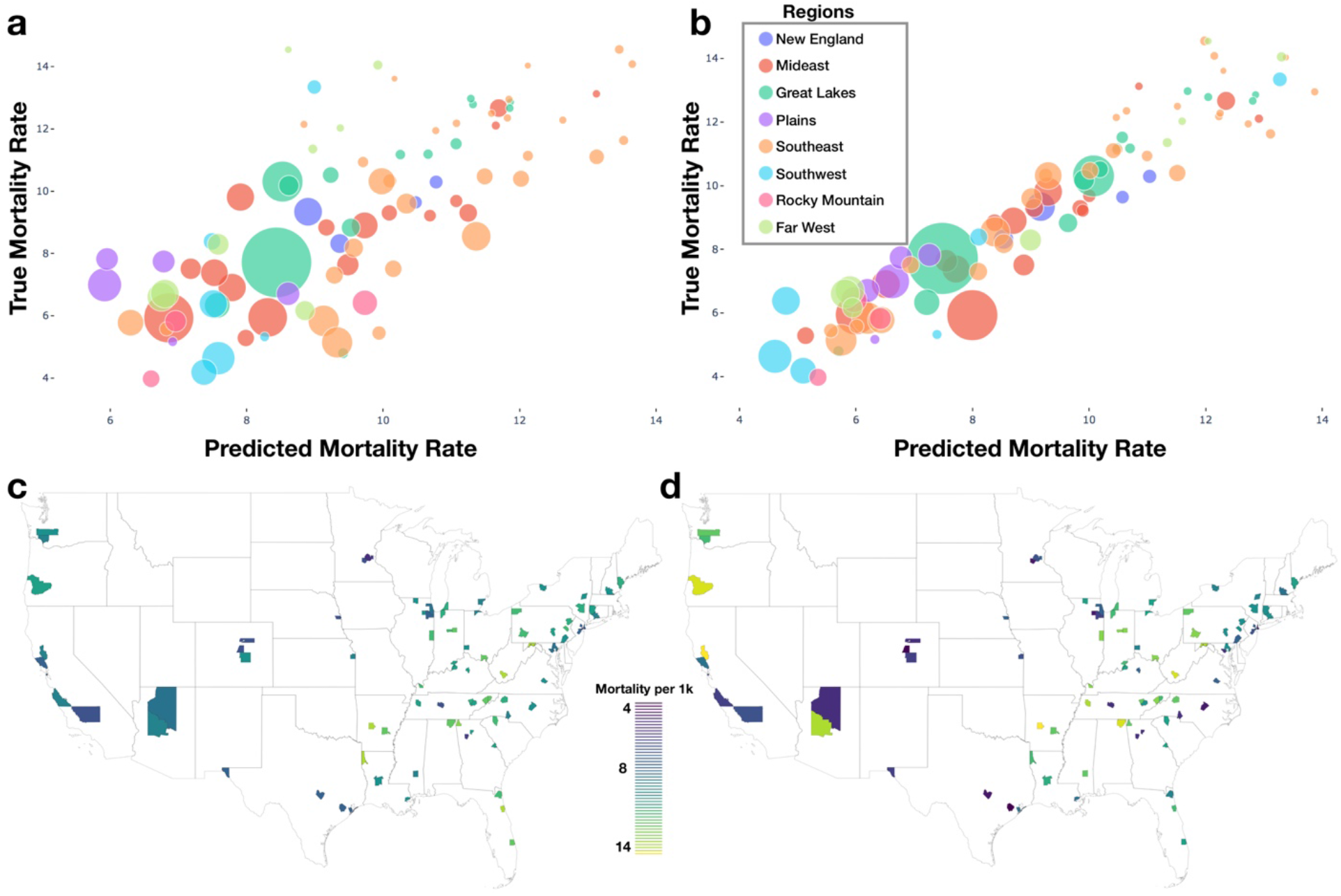
Model Results: a) deep learning predicted versus true mortality for each test county; b) linear model predicted versus true mortality for each test county; a-b) legend contains look-up dictionary for naming of US regions featured in the rest of the study; size of bubble is correspondent to population size of county; c) predicted mortality plotted geographically for test counties; d) true mortality plotted geographically for test counties

### Model Interpretation Results

Using SHAP to identify relevant features in satellite images across these test counties, we noticed that the model was able to associate common features associated with the socioeconomic status of that community to reductions in mortality. Generally, we were able to spot instances in which sidewalks, driveways, curved roads, hiking trails, baseball fields and light-colored roofs were associated with reductions in mortality (Figure 1d, 4). Conversely, we noticed instances where centerlines of large roads and shadows of buildings and trees were positively associated with mortality. To corroborate the evidence found using SHAP, we plotted small images representing patterns that the first convolutional layer had learned and then convolved each of these patterns with three select images (Supplementary Figures 3-6). Across three images from California, Virginia, and Georgia, filters such as 12, 29 and 43 focused on bright cues and were able to highlight driveways, sidewalks, walkways and baseball fields, while filters 13 and 46 were able to pick up on contours and nuanced shape-based patterns.

**Figure 4:**
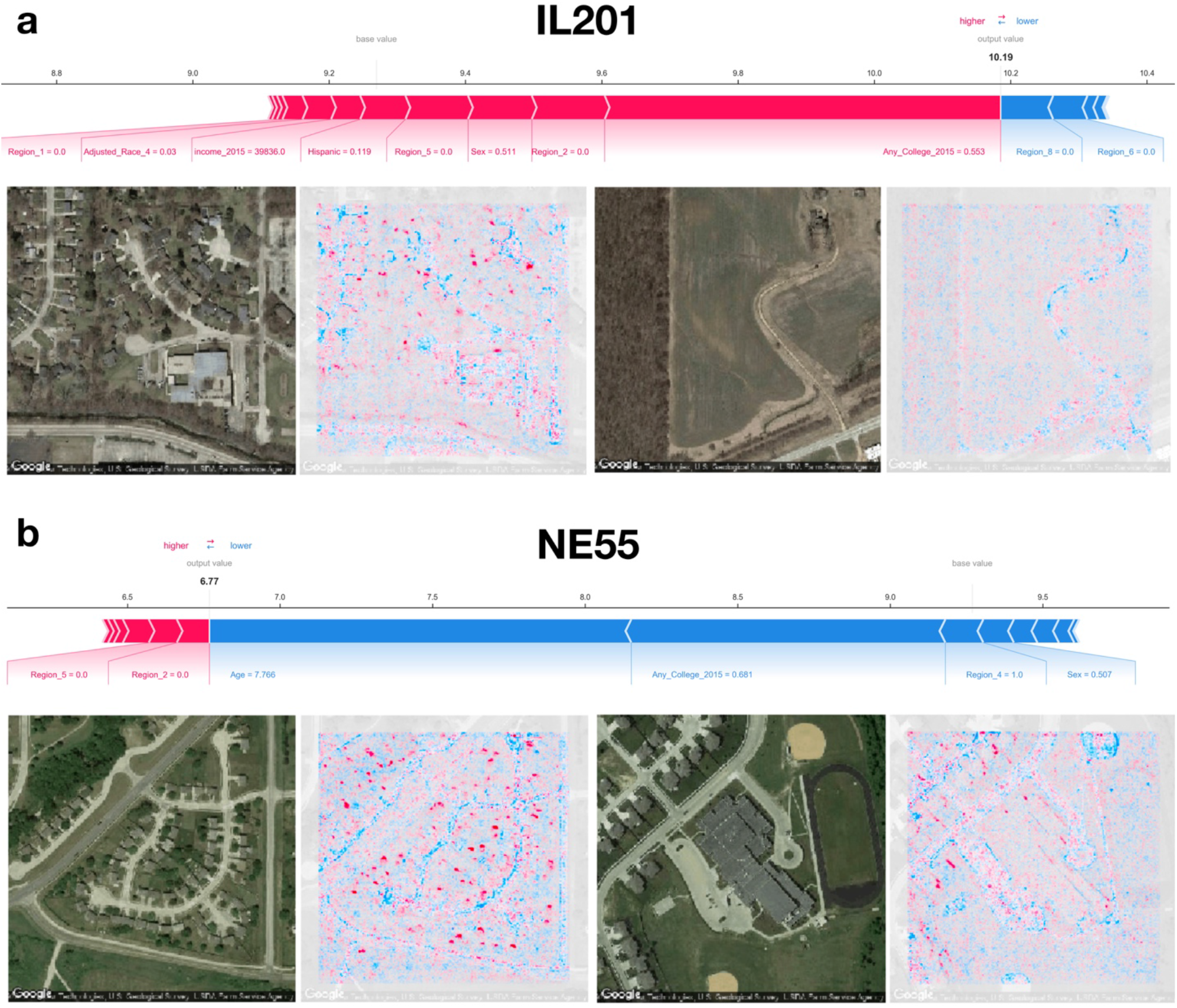
SHAP Image Interpretations: a-b) Shapley features extracted from both covariate and image models for select images from counties in Illinois and Nebraska (FIPS codes IL201 and NE55 respectively); blue coloring indicates features associated with reductions in mortality, red indicates association with increased mortality; additional image interpretations can be found in Figure 1d

From the county-level covariate linear model, we observed a reduction in 13 deaths per 100,000 individuals for an 1% increase in the proportion of those who attend college (regression coefficient *β* = − 12.6± 0.8), and increase of 214 deaths per 100,000 individuals for 5-year increases in average population age (*β* = 2.1±0.1); these were the most important predictors of county mortality (Figure 5a-c, Supplementary Table 3-6). Increased Hispanic (*β* = −6.6±0.5), female (*β* =42.5±6.4), or Asian race proportions (*β* = −7.8± 1.0), and living in the western United States (*β* = −1.1± 0.3) as compared to the other regions, were found to be protective county-level factors against mortality (Figure 3a-b, Supplementary Tables 2-4). County-level covariate mortality predictors are ranked in order of decreasing importance in Figure 5a-b and Supplementary Tables 3-6.

**Figure 5:**
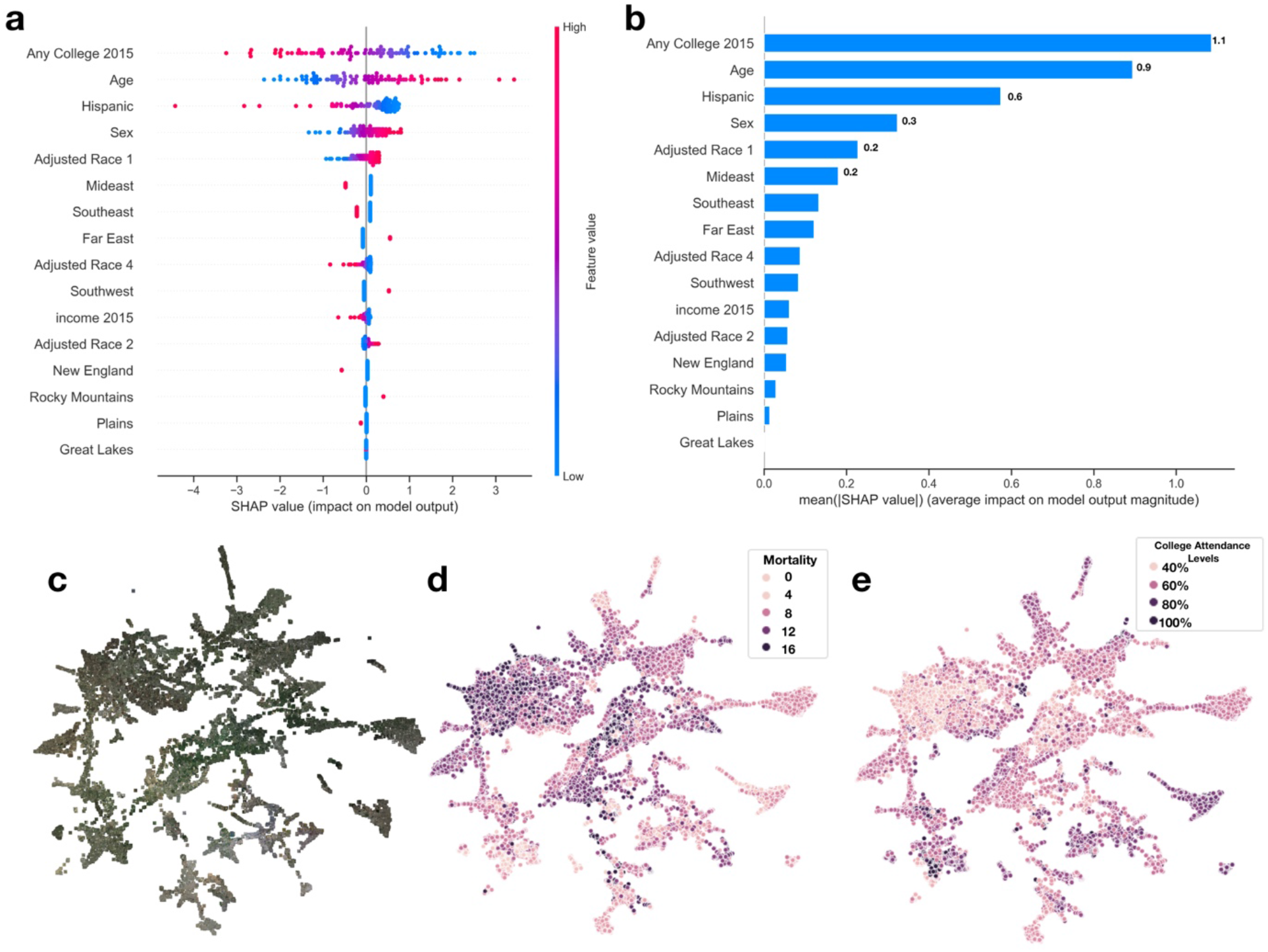
SHAP Summary of Covariate Predictors and Image Embeddings. a) SHAP summary plot for covariate model evaluated on test set, covariates ranked by feature importance; b) SHAP rankings of overall importance of covariates; c-e) 2-D UMAP plot of image features derived from images using deep learning model: c) actual images overlaying 2-D coordinates; d) colored by true county mortality; e) colored by average college attendance; we note here that our model has been explicitly trained to associate image features with mortality; the separability of college attendance over the image features is an artifact of the association between education status and mortality

We identified demographic and mortality-related sources of significant variation between the ten image feature clusters (Figure 5c-d, 6a) established using UMAP and Spectral Clustering (Figures 6b, Supplementary Table 7). Images in cluster 7 were associated with the highest mortality rate of all of the clusters (mortality rate of 11 deaths per 1,000 individuals) and generally included counties from the southeastern United States (Supplementary Figure 7). Income and educational status were lower on average compared to the other clusters. We contrast cluster 7 with cluster 2, a small cluster with the third lowest mortality rates (6.6 deaths per 1,000 individuals) of the 10 clusters (Figure 6; Supplementary Figures 8-10,11; Supplementary Table 7) and identified variation within cluster 5; we include a brief discussion in the supplementary materials. Many of the covariate mortality predictors across the test set demonstrated strong associations with the deep learning predicted county-level mortality rates (Supplementary Table 8).

**Figure 6:**
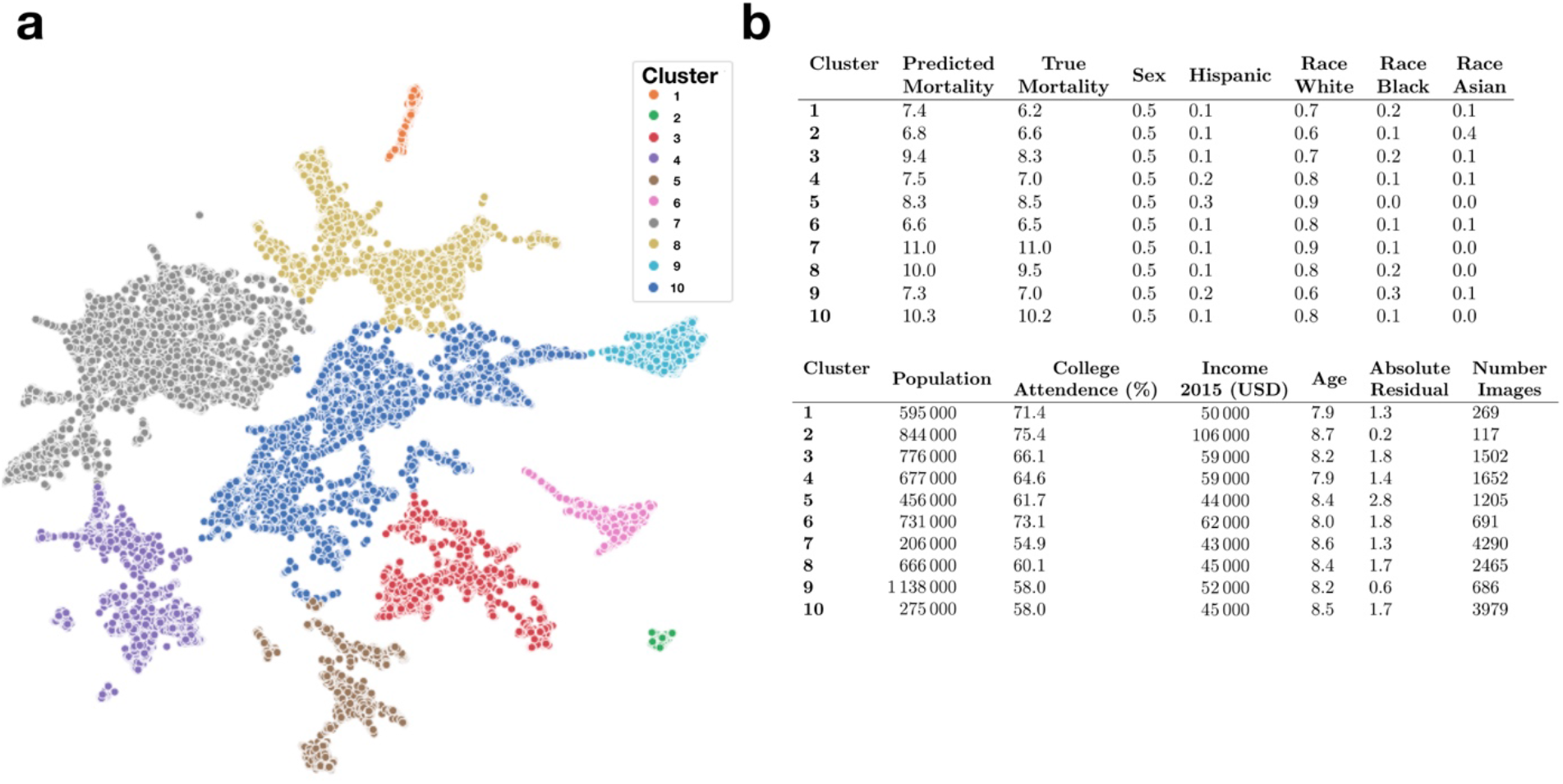
Key Covariate Characteristics of Found Image Clusters: a) 2-D UMAP plots colored by cluster as estimated through Spectral Clustering; b) averaged covariate statistics for each image cluster; average population and income estimates rounded to thousands place; statistics average county-level covariates, weighted by constituent images, covariates include: 1) average predicted mortality for the cluster, 2) average true mortality assigned to the cluster, 3) proportion female/male, 4) proportion Hispanic, 5) proportion white, black or asian, 6) average population, 7) proportion attend college, 8) average household income, 9) average age of population (in increments of 5-years per description in the supplementary materials; e.g., 7.9*5 years=39.5 years old), 10) Absolute Residual between true and predicted mortality, 11) number of images for cluster / size of cluster

## Discussion

Here, we demonstrated the feasibility of using deep learning and satellite imagery to predict county mortality across the United States, extending methods and approaches from prior deep learning studies that linked the built environment and health-related factors. Furthermore, we established clusters of images that represented various demographic groups and interrogate these learned patterns to find additional corroborating evidence.

While the covariate model significantly outperformed our deep learning model and identified important predictors that corroborated with prior literature, the deep learning approach identified meaningful built features of the environment, representing a benchmark for performance from which to compare future applications [39, 40].

We only sampled neighborhood characteristics found around schools. For instance, there is likely a greater number of sports fields and playgrounds surrounding schools than in random locations in the county. In many of the neighborhoods, regardless of mortality rates, there is likely more green space surrounding schools. While prior literature suggests green space is associated with positive health factors [41], the fact that this green space is nearly ubiquitous in neighborhoods near these schools may cause our model to down-weight these urban design factors. It also appeared that predictive performance increased by sampling the surrounding neighborhood around each school, suggesting that the surrounding neighborhood contains more important information associated with mortality than utilizing the schools alone.

The evidence of what our deep learning model found to be indicative of mortality can be corroborated with existing literature on associations of these image features with higher socioeconomic status, decreased obesity, and greater designs in urban planning [41–43]. For instance, we found that sidewalks, driveways, curvy roads, hiking trails and baseball fields, amongst other factors, were related to lower mortality; these factors were also uncovered from inspection of learned convolutional filters (Supplementary Figures 3-6).

The assigned importance given to image features such as baseball fields does not imply that these components are necessary for urban design but does further elucidate a suite of modifiable community factors to design interventions. For instance, accessibility to trails, while indicative of a county with high socioeconomic status [44, 45], provides a convenient means to exercise but can also provide access to other portions of a community, allowing more social mobility and access to health care [46, 47].

Despite our strong findings, there were some limitations to our study. The design of this study was ecological with sampling at one time point for both images and mortality (cross-sectional). This precluded our ability to show temporality and that the built environment was causally associated with mortality. The images also were taken in 2018 while we assessed mortality in 2015. While we do not expect areas surrounding schools to change substantially, we cannot confirm the extent of land-use alterations between 2015 and 2018. We also acknowledge the shortfalls associated with utilizing counties as the primary spatial unit of analysis in this study. While county-level demographic and mortality information are more widely and freely available, mortality rates and demographic factors can vary widely within counties; thus, counties may not always represent the best unit of analysis for capturing the complete heterogeneity in population demographics and mortality rates. In future work, longitudinal image monitoring could be used to forecast increases in mortality rate and help determine optimal intervention strategies that complement county/state planning and development efforts. We assessed county mortality rates using a small percentage of schools from the county, making generalizations to counties included in the study and counties not in our dataset. We were able to predict crude mortality rates accurately within a subsample of these counties. However, our primary modeling approach could not delineate how much the corresponding model-identifiable neighborhood features were intermediates reflective of mortality associated demographics or were directly associated with mortality. Additionally, the deep learning model that combined county-level demographic and satellite imagery data did not surpass the performance of the demographic-only model. We incorporated county-level demographic data into images that have varying degrees of importance for the prediction of mortality; we suspect that adjusting and including the images that are less predictive of mortality may partially explain the performance of the demographic plus satellite image model.

Higher correspondence between predicted and true mortality may be achieved by increasing the number of schools selected but tapers off with a larger sampling area, which is also suggestive that satellite images beyond a certain distance from a school are not as predictive of mortality. In addition, we may be able to identify more ubiquitous points of interest other than schools from which to sample. While schools were selected as a proxy for residential neighborhoods (prior research on impact of surrounding neighborhood of schools on health disparities), inclusion and exclusion of other points of interest (e.g., proximity to fast-food establishments) may result in more accurate models, which can potentially allow for in-depth hypotheses testing of the impact of urban planning on mortality and health disparities. Another opportunity is to integrate neighborhood land use information (e.g., residential/commercial) into the selection of images and adjustments during the modeling approach, though such approaches require accurate up-to-date mapping information [48–55].

While we acknowledge prior literature documenting the potential for shadow effects to confound aerial imagery analysis, we also note that our model was able to pick up on key factors associated with mortality by utilizing learned filters with shape, color and intensity [56, 57] (Supplementary Figures 1-4). Possible removal or augmentation of these shadows may cause the model to focus on other important characteristics pertinent to higher mortality prediction [56– 58]. Finally, although we sampled counties randomly, our sampling scheme preferred populous counties, and had assumed that these counties would solely contain suburban and residential land-use patterns. However, even populous counties contained rural areas that may have obfuscated our ability to sample ubiquitous land-use regions for the mortality prediction potentially biasing the results. Recent deep learning works have focused on grasping health factors such as access to care in the rural setting [59] and could be employed in the context of mortality in the future.

The deep learning approaches used were able to achieve remarkable performance given technical challenges associated with sampling images over large geographic regions. Nor were covariate measures or temporality incorporated into the model. Different sampling techniques [60], feature aggregation measures, evaluating or producing higher resolution images [61], direct estimation of neighborhood demographic factors, and segmentation of various land-use objects can potentially provide more accurate and interpretable models for studying health and disease [24, 62, 63]. The images may have also been sampled during various points throughout the year, thus results may have been affected by seasonality, however, it is likely that the images had been collected randomly with respect to seasonality and the fact that the model was able to distinguish mortality rates further attests of the ability to learn features less tied to seasonality. Future applications could include deep learning methodologies that are able to account for effects of seasonality [64, 65]. Transfer learning from other GIS-corroborated data may also improve the model’s performance. Regardless, our approach is scalable and uses open access data enabling further exploration.

While the county-level covariate prediction model obtained higher accuracy versus the deep learning model, a major limitation of the county-covariate approach pertains to evolving guidelines on the reporting of sex and race in public health research studies. County-level demographic characteristics were extracted from the Surveillance, Epidemiology, and Ends Results (SEER) Program, yet numerous publications have critiqued this reporting system for failing to incorporate sexual and gender minorities (e.g., gradations of sex identification, non-binary sex, LGBTQ+) and racial minority groups (e.g., description of individuals based on regional descent). Some of the criticisms of such reporting standards are that they enforce socially defined racial, ethnic, and gender divisions/constructs which further separates such groups (potentially contributing to health disparities they seek to study and reduce), disregards self-reported identification that transcends social structure imbued by support of such classifications, and reduces the potential to study additional meaningful health disparities between these minority groups (e.g., delineating health outcomes for individuals of Southeast Asian descent versus that of Asian Americans as a whole). These issues have made it difficult to study and document health outcomes for these minority groups. Conversely, some have pointed out that these reporting standards have still proven useful for studying health disparities (e.g., allostatic load and stress amongst minority groups and effect of segregation on health access) in order to devise policies to alleviate these differences and that further subclassification may make it more difficult to assess meaningful differences. In response, standards have shifted towards asking multiple questions in demographic surveys which provide further clarification on self-identity and country of origin in addition to these coarse measures of sex and race. Conversations around the inclusion of sex and race in medical research studies are especially pertinent given persistent violence against minority groups that have more recently prompted a national conversation on such issues. In accordance with 2021 reporting standards on race and sex, we acknowledge the limitations of our findings with respect to these issues (using broad racial groups and binary sex identification), as such limitations reflect 2015 survey standards. While it is outside of the study scope to modify the analysis in response to data limitations, we have included a number of citations for the reader to explore facets of this important issue as part of a larger conversation on the role of sex and race in public health research studies [66–84].

Despite the study limitations, this approach to assess mortality using CNNs has much external applicability and room for improvement. Offshoots of this approach may further explore how the built environment affects mortality in more precise estimates, such as cities, or explore rural areas exclusively [85]. Additionally, our approach demonstrates that built image features of the environment are correlated with demographic characteristics. Future mortality research that lacks the ability to attain particular covariate information due to feasibility or expense costs, could thus infer mortality using images alone or in combination with some known covariates. Applications beyond assessing mortality as an outcome could benefit from this approach and could use other landmarks as a sampling strategy for prediction instead of schools. Sampling from landmarks such as schools reduces the stochastic selection of images from large areas and potentially obviate the need to select and download huge swaths of images. However, if the researcher is able to overcome the resources required to download all of the images from within each county, future methodological advances should consider methods to assign importance scores to each satellite image or weight each county by their perceived macro-level importance for improved mortality prediction. Combining such methods with demographic information and contextualizing satellite images by neighboring images may yield models that ignore these demographic community factors. Ultimately, these tools could then be used by epidemiologists and policy makers to identify clusters of exposures or diseases such as cancer, arsenic exposure, or infectious diseases for more targeted interventions [24].

## Conclusion

We found that mortality could be predicted from satellite imagery. Future use of deep learning and satellite imagery may assist in forming targeted public health interventions and policy changes.

## Data Availability

Mortality death certificate data can be downloaded from CDC Wonder, per third-party request, available at the following URL: https://www.cdc.gov/nchs/data_access/cmf.htm. Google Maps Satellite Images are publicly available and can be downloaded using the Google Static Maps API.

https://github.com/jlevy44/SatelliteCountyMortalityPrediction

## Author Contributions

Concept and design: JJL, RML, and TAM

Acquisition, analysis, or interpretation of the data: All authors

Drafting of the manuscript: JJL, RML, and TAM

Critical revision to the manuscript for important intellectual content: All authors

Statistical analysis: JJL, RML, TAM

Administrative, technical, or material support: JJL, RML and TAM Supervision: TAM

## Acknowledgements

The authors would like to thank Christopher L. Henry and Jorge

F. Lima for discussion of image interpretations.

## Availability of data and materials

Mortality death certificate data can be downloaded from CDC Wonder, per third-party request, available at the following URL: https://www.cdc.gov/nchs/data_access/cmf.htm. Google Maps Satellite Images are publicly available and can be downloaded using the Google Static Maps API. Analysis code can be found at the following GitHub URL: https://github.com/jlevy44/SatelliteCountyMortalityPrediction.

## Ethics approval and consent to participate

Not applicable.

## Consent for Publication

Not applicable.

