## Supplementary Materials for "Using Satellite Images and Deep Learning to Identify Associations Between County-Level Mortality and Residential Neighborhood Features Proximal to Schools: A Cross-Sectional Study"

### Supplementary Material

#### Dataset and Code Availability

The code used to acquire and analyze the dataset and results can be found in the following GitHub repository: https://github.com/jlevy44/SatelliteCountyMortalityPrediction . Data for county-level covariate information is not available from our repository as permissions to access the data source are required.

#### Availability of High-Resolution Materials

Due to large file size, high-resolution manuscript figures and ultra-high-resolution image of UMAP image embeddings overlaid with satellite images, and additional SHAP image plots are available upon request or at the aforementioned GitHub repository.

#### More Information on Selection of Variables for Covariate Model

For adjusted linear models, age, sex, and Hispanic status, were extracted from the death certificates. Age was divided into 19 categories brackets and race was coded as White, Black, and Asian or Pacific Islander. Individuals considered Alaskan or Native American were removed prior to mortality calculations due to inconsistent coding of race between the CDC Wonder and SEER datasets. Percentage of college attainment was imputed for counties without information as the median value for all available counties. Median per capita county income and the median value for all available counties was also imputed for counties with any missing per capita income. Public school information was collected from the National Center for Education Statistics. Covariate information was collected from the SEER program database.

Ages were binned into 5-year brackets and thus turned into an ordinal variable. The deaths were averaged across the brackets to yield a final death estimate. Race was changed into a ratio where the number of deaths of individuals in each county of a certain race were divided by the total number of individuals in each county creating a race ratio. Percentages of deaths were calculated by sex and Hispanic status. Median income and any college were not changed. Grouping regions include: New England, Mideast, Great Lakes, Plains, Southeast, Southwest, Rocky Mountain, and Far West.

**Supplementary Table 1**: **Average county characteristics by data set**; 2 counties in the training set did not have income data, values were imputed by region; Averaged county mortality rates were not weighted by population size

| | **Training** | **Testing** | **Validation** |
|---|---|---|---|
| *n (%)* | 279 (65) | 86 (20) | 65 (15) |
| *Mean (SD) for all counties* | | | |
| **Population (in 1000s)** | 537 (823) | 491 (677) | 505 (555) |
| **Mortality Rate (per 1000 people)** | 9.29 (2.8) | 9.21 (2.8) | 9.35 (2.9) |
| **Per capita income ($)*** | 47000 (13000) | 48900 (17400) | 47500 (11800) |
| **Any College Rate (%)** | 60.0 (9.6) | 60.4 (10.4) | 60.8 (10.3) |

**Supplementary Table 2:** Counties selected for training, validation and testing sets

| DATASET | COUNTIES |
|---|---|
| **TRAINING** | AL101,AL121,AL15,AL3,AL33,AL43,AL49,AL55,AL73,AL77,AL89,AL95,AL97,AR119,AZ13,AZ19,AZ21,CA1,CA107,CA13,CA19,CA31,CA37,CA59,CA61,CA65,CA67,CA7,CA71,CA81,CA83,CA89,CA95,CO123,CO31,CO59,CO69,CT1,DC1,DE3,DE5,FL1,FL101,FL103,FL105,FL107,FL115,FL119,FL127,FL15,FL21,FL5,FL53,FL57,FL69,FL71,FL73,FL81,FL83,FL85,FL86,FL9,FL91,FL95,FL99,GA121,GA135,GA21,GA215,GA57,GA63,GA89,IA103,IA153,IA169,IL111,IL115,IL119,IL143,IL163,IL167,IL179,IL43,IL89,IL95,IL99,IN163,IN177,IN3,IN57,IN89,IN95,IN97,KY199,LA19,LA33,LA51,LA97,MA1,MA13,MA17,MA21,MA23,MA25,MA27,MA5,MA9,MD13,MD3,MD510,ME3,ME5,MI125,MI145,MI147,MI17,MI25,MI49,MI65,MI81,MI99,MN139,MN163,MN3,MN37,MO183,MO510,MO95,MS47,MT31,NC125,NC157,NC159,NC161,NC179,NC19,NC21,NC35,NC51,NC57,NC67,NC71,NC81,NC89,NE109,NE153,NH15,NJ17,NJ21,NJ25,NJ27,NJ29,NJ3,NJ31,NJ35,NJ39,NJ5,NJ7,NJ9,NM1,NV3,NV510,NY103,NY119,NY13,NY15,NY29,NY5,NY59,NY63,NY65,NY7,NY81,NY87,OH13,OH139,OH155,OH167,OH29,OH35,OH41,OH43,OH49,OH61,OH7,OH81,OH85,OH89,OH93,OK109,OK37,OR11,OR29,OR39,OR47,OR5,OR51,OR67,PA101,PA111,PA13,PA133,PA17,PA19,PA21,PA25,PA3,PA41,PA45,PA49,PA5,PA51,PA69,PA7,PA71,PA73,PA79,PA81,PA85,PA91,PA95,SC19,SC31,SC45,SC51,SC59,SC7,SC73,SC79,SD83,TN149,TN163,TN35,TN65,TX113,TX121,TX181,TX201,TX209,TX213,TX215,TX221,TX245,TX265,TX27,TX29,TX339,TX39,TX41,TX439,TX479,TX61,UT11,UT35,UT49,VA13,VA153,VA179,VA510,VA59,VA770,VA810,VA87,WA11,WA21,WA61,WA63,WI133,WI135,WI25,WI79,WV11,WV33,WV39,WV55,WY5 |
| **VALIDATION** | AL103,AZ15,CA111,CA73,CA77,CA85,CA99,CO13,CO5,CT3,FL11,FL117,FL33,FL55,FL61,FL97,IA49,ID1,IL1,IL97,IN141,KS161,KS173,KY195,MA3,MD31,MD33,ME11,MI161,MN137,MO189,MO77,NC171,NC45,NJ1,NV31,NY27,NY67,NY71,OH113,OH151,OH153,OH23,OH87,OH99,OK143,OR33,PA107,PA11,PA125,PA97,RI7,TN145,TN157,TN37,TX355,TX423,TX491,TX85,UT5,WA33,WA53,WA9,WI101,WV107 |
| **TEST** | AL71,AR51,AR69,AZ25,AZ5,CA29,CA33,CA53,CA75,CA97,CO1,CO35,CO41,CT5,CT9,FL111,FL31,FL35,GA117,GA245,GA295,GA67,IL183,IL197,IL201,IL31,IL93,IN19,IN53,IN67,IN91,KS91,KY111,KY145,KY67,LA17,LA71,LA79,MD1,MD27,MD5,ME31,MI163,MI21,MN123,MN19,MN53,MS67,NC119,NC183,NC193,NE55,NH11,NJ13,NJ23,NY1,NY111,NY47,NY55,NY61,NY85,OH145,OH17,OH95,OR19,PA129,PA29,PA39,PA43,PA77,PA89,SC83,TN1,TN179,TN187,TN53,TN59,TN93,TX141,TX157,TX167,TX453,VA41,WA15,WA41,WV81 |

**Supplementary Table 3**: Standardized Coefficients of Linear Regression Model

| | coef | std err | t | $P(T>\|t\|)$ | [0.025 | 0.975] |
|---|---|---|---|---|---|---|
| **Constant** | 9.39 | 0.05 | 192.32 | 0.00 | 9.29 | 9.48 |
| **Age** | 1.46 | 0.07 | 20.32 | 0.00 | 1.32 | 1.60 |
| **Any College 2015** | −1.24 | 0.08 | −15.79 | 0.00 | −1.39 | −1.08 |
| **Hispanic** | −0.95 | 0.07 | −14.23 | 0.00 | −1.09 | −0.82 |
| **Proportion Male** | 0.43 | 0.06 | 6.67 | 0.00 | 0.30 | 0.56 |
| **Far West** | 0.30 | 0.05 | 5.67 | 0.00 | 0.20 | 0.41 |
| **Southwest** | 0.26 | 0.05 | 5.09 | 0.00 | 0.16 | 0.36 |
| **Proportion Asian** | −0.22 | 0.06 | −3.77 | 0.00 | −0.34 | −0.11 |
| **Income 2015** | −0.20 | 0.08 | −2.64 | 0.01 | −0.35 | −0.05 |
| **Mideast** | −0.19 | 0.05 | −4.22 | 0.00 | −0.28 | −0.10 |
| **Southeast** | −0.15 | 0.04 | −3.28 | 0.00 | −0.23 | −0.06 |
| **New England** | −0.12 | 0.05 | −2.41 | 0.02 | −0.21 | −0.02 |
| **Proportion White** | 0.12 | 0.03 | 3.54 | 0.00 | 0.05 | 0.18 |
| **Plains** | −0.06 | 0.05 | −1.19 | 0.24 | −0.16 | 0.04 |
| **Rocky Mountains** | 0.02 | 0.05 | 0.47 | 0.64 | −0.08 | 0.13 |
| **Proportion Black** | −0.02 | 0.04 | −0.45 | 0.65 | −0.10 | 0.07 |
| **Great Lakes** | 0.01 | 0.04 | 0.17 | 0.86 | −0.08 | 0.09 |

**Supplementary Table 4**: Non-Standardized Coefficients of Linear Regression Model

| | coef | std err | t | $P(T>\|t\|)$ | [0.025 | 0.975] |
|---|---|---|---|---|---|---|
| **Constant** | −15.5 | 2.2 | −7.1 | 0.0 | −19.7 | −11.2 |
| **Proportion White** | −3.4 | 0.7 | −4.6 | 0.0 | −4.8 | −1.9 |
| **Proportion Black** | −4.3 | 1.0 | −4.5 | 0.0 | −6.2 | −2.5 |
| **Proportion Asian** | −7.8 | 1.0 | −7.7 | 0.0 | −9.7 | −5.8 |
| **Hispanic** | −6.6 | 0.5 | −14.2 | 0.0 | −7.5 | −5.7 |
| **Proportion Male** | 42.5 | 6.4 | 6.7 | 0.0 | 30.0 | 55.1 |
| **Mean Age** | 2.1 | 0.1 | 20.3 | 0.0 | 1.9 | 2.3 |
| **Any College 2015** | −12.6 | 0.8 | −15.8 | 0.0 | −14.1 | −11.0 |
| **New England** | −2.5 | 0.4 | −6.7 | 0.0 | −3.3 | −1.8 |
| **Mideast** | −2.5 | 0.3 | −8.0 | 0.0 | −3.1 | −1.9 |
| **Great Lakes** | −2.0 | 0.3 | −6.3 | 0.0 | −2.6 | −1.4 |
| **Plains** | −2.2 | 0.3 | −6.8 | 0.0 | −2.9 | −1.6 |
| **Southeast** | −2.3 | 0.3 | −7.2 | 0.0 | −2.9 | −1.7 |
| **Rocky Mountains** | −1.8 | 0.3 | −5.4 | 0.0 | −2.5 | −1.2 |
| **Far West** | −1.1 | 0.3 | −3.7 | 0.0 | −1.6 | −0.5 |
| **Southwest** | −1.1 | 0.3 | −3.3 | 0.0 | −1.7 | −0.4 |
| **Income 2015** | $-1.6 \cdot 10^{-05}$ | $5.9 \cdot 10^{-06}$ | −2.6 | 0.0 | $-2.7 \cdot 10^{-05}$ | $-4.0 \cdot 10^{-06}$ |

**Supplementary Table 5**: Fit Statistics for Linear Regression Model on Training and Validation Set

| | |
|---|---|
| **R-squared:** | 0.904 |
| **Adj. R-squared:** | 0.900 |
| **F-statistic:** | 221.2 |
| **Prob (F-statistic):** | 7.07E-158 |
| **Log-Likelihood:** | -446.16 |
| **AIC:** | 922.3 |
| **BIC:** | 979.9 |

**Supplementary Table 6**: SHAP Feature Importance Scores over Test Set

| | |
|---|---|
| **Any College 2015** | 1.1 |
| **Mean Age** | 0.9 |
| **Hispanic** | 0.6 |
| **Proportion Male** | 0.3 |
| **Proportion White** | 0.2 |
| **Mideast** | 0.2 |
| **Proportion Black** | 0.1 |
| **Proportion Asian** | 0.1 |
| **Income 2015** | 0.1 |
| **New England** | 0.1 |
| **Southeast** | 0.1 |
| **Southwest** | 0.1 |
| **Far West** | 0.1 |
| **Great Lakes** | 0.0 |
| **Plains** | 0.0 |
| **Rocky Mountains** | 0.0 |

**Supplementary Table 7**: Weighted T-Test Comparison between Clusters for Differences in Key Covariates: a) Mortality, b) Income, c) Hispanic, d) Any College, e) Age:

| a | 1 | 2 | 3 | 4 | 5 | 6 | 7 | 8 | 9 | 10 |
|---|---|---|---|---|---|---|---|---|---|---|
| 1 | | 2E-01 | 1E-40 | 3E-07 | 1E-21 | 3E-01 | 4E-194 | 4E-103 | 3E-20 | 1E-121 |
| 2 | | | 2E-13 | 5E-01 | 4E-07 | 1E+00 | 5E-78 | 2E-39 | 7E-07 | 2E-46 |
| 3 | | | | 2E-50 | 1E+00 | 2E-75 | 6E-278 | 1E-58 | 3E-43 | 1E-126 |

| | | | | | | | | | | |
|---|---|---|---|---|---|---|---|---|---|---|
| **4** | | | | | 5E-37 | 5E-09 | 0E+00 | 7E-228 | 1E+00 | 0E+00 |
| **5** | | | | | | 4E-42 | 4E-167 | 3E-26 | 1E-24 | 2E-70 |
| **6** | | | | | | | 0E+00 | 2E-207 | 2E-18 | 8E-256 |
| **7** | | | | | | | | 8E-125 | 0E+00 | 9E-51 |
| **8** | | | | | | | | | 4E-156 | 1E-21 |
| **9** | | | | | | | | | | 2E-198 |
| **10** | | | | | | | | | | |

| b | 1 | 2 | 3 | 4 | 5 | 6 | 7 | 8 | 9 | 10 |
|---|---|---|---|---|---|---|---|---|---|---|
| **1** | | 1E-235 | 2E-05 | 4E-05 | 2E-24 | 1E-25 | 4E-32 | 2E-16 | 4E-05 | 4E-12 |
| **2** | | | 3E-69 | 1E-69 | 0E+00 | 1E-117 | 0E+00 | 0E+00 | 0E+00 | 0E+00 |
| **3** | | | | 1E+00 | 1E-64 | 2E-01 | 1E-210 | 6E-101 | 5E-08 | 2E-138 |
| **4** | | | | | 1E-63 | 2E-01 | 1E-211 | 1E-99 | 4E-07 | 4E-139 |
| **5** | | | | | | 1E-157 | 6E-04 | 9E-03 | 9E-85 | 1E+00 |
| **6** | | | | | | | 0E+00 | 3E-213 | 1E-40 | 2E-206 |
| **7** | | | | | | | | 4E-26 | 1E-118 | 1E-18 |
| **8** | | | | | | | | | 5E-71 | 1E+00 |
| **9** | | | | | | | | | | 5E-54 |
| **10** | | | | | | | | | | |

| c | 1 | 2 | 3 | 4 | 5 | 6 | 7 | 8 | 9 | 10 |
|---|---|---|---|---|---|---|---|---|---|---|
| **1** | | 2E-31 | 4E-01 | 2E-17 | 4E-47 | 1E+00 | 1E-147 | 3E-06 | 1E-274 | 1E-03 |
| **2** | | | 5E-09 | 7E-03 | 1E-15 | 3E-05 | 1E-171 | 2E-20 | 2E-101 | 3E-12 |
| **3** | | | | 1E-90 | 4E-223 | 1E+00 | 9E-243 | 2E-07 | 1E-184 | 2E-05 |
| **4** | | | | | 1E-75 | 8E-43 | 0E+00 | 2E-179 | 1E+00 | 2E-217 |
| **5** | | | | | | 5E-110 | 0E+00 | 0E+00 | 5E-49 | 0E+00 |
| **6** | | | | | | | 7E-149 | 2E-05 | 7E-109 | 8E-04 |
| **7** | | | | | | | | 8E-207 | 0E+00 | 2E-174 |
| **8** | | | | | | | | | 1E-319 | 1E+00 |
| **9** | | | | | | | | | | 1E-221 |
| **10** | | | | | | | | | | |

| d | 1 | 2 | 3 | 4 | 5 | 6 | 7 | 8 | 9 | 10 |
|---|---|---|---|---|---|---|---|---|---|---|
| **1** | | 5E-13 | 7E-17 | 2E-17 | 8E-64 | 4E-06 | 3E-139 | 5E-100 | 8E-259 | 1E-116 |
| **2** | | | 4E-23 | 2E-19 | 4E-59 | 8E-07 | 1E-96 | 1E-83 | 3E-291 | 3E-88 |
| **3** | | | | 6E-03 | 7E-33 | 1E-69 | 2E-264 | 4E-90 | 5E-90 | 4E-167 |
| **4** | | | | | 2E-11 | 1E-65 | 2E-191 | 6E-43 | 2E-40 | 5E-103 |
| **5** | | | | | | 1E-188 | 5E-91 | 2E-07 | 6E-25 | 7E-33 |
| **6** | | | | | | | 0E+00 | 3E-290 | 0E+00 | 0E+00 |
| **7** | | | | | | | | 1E-96 | 7E-14 | 1E-44 |

| | | | | | | | | | | |
|---|---|---|---|---|---|---|---|---|---|---|
| **8** | | | | | | | | | 5E-10 | 3E-19 |
| **9** | | | | | | | | | | 1E+00 |
| **10** | | | | | | | | | | |

| e | 1 | 2 | 3 | 4 | 5 | 6 | 7 | 8 | 9 | 10 |
|---|---|---|---|---|---|---|---|---|---|---|
| **1** | | 2E-104 | 2E-31 | 1E+00 | 2E-14 | 1E+00 | 7E-153 | 8E-43 | 1E-41 | 1E-89 |
| **2** | | | 2E-30 | 2E-87 | 3E-02 | 1E-157 | 1E+00 | 7E-05 | 5E-102 | 2E-04 |
| **3** | | | | 6E-113 | 3E-08 | 5E-60 | 2E-218 | 6E-29 | 4E-04 | 5E-84 |
| **4** | | | | | 7E-78 | 2E-03 | 0E+00 | 2E-208 | 2E-56 | 0E+00 |
| **5** | | | | | | 3E-31 | 2E-37 | 1E+00 | 7E-09 | 4E-05 |
| **6** | | | | | | | 0E+00 | 5E-92 | 3E-60 | 2E-192 |
| **7** | | | | | | | | 2E-69 | 3E-172 | 8E-56 |
| **8** | | | | | | | | | 5E-30 | 2E-05 |
| **9** | | | | | | | | | | 4E-77 |
| **10** | | | | | | | | | | |

**Supplementary Table 8**: Goodness-of-fit for univariable regression of predicted mortality to each of the mortality covariate predictors to directly associate image features with mortality factors

| Covariate | Adjusted R-Squared | P-Value |
|---|---|---|
| **Mean Age** | 0.971 | **4.35E-67** |
| **Proportion Male** | 0.961 | **8.55E-62** |
| **Proportion White** | 0.944 | **4.33E-55** |
| **Any College 2015** | 0.894 | **1.85E-43** |
| **Income 2015** | 0.797 | **2.29E-31** |
| **Proportion Black** | 0.513 | **3.92E-15** |
| **Southeast** | 0.402 | **2.62E-11** |
| **Hispanic** | 0.341 | **1.78E-09** |
| **Proportion Asian** | 0.293 | **3.65E-08** |
| **Mideast** | 0.177 | **2.95E-05** |
| **Great Lakes** | 0.160 | **7.51E-05** |
| **Far West** | 0.057 | **1.47E-02** |
| **New England** | 0.036 | **4.26E-02** |
| **Southwest** | 0.034 | **4.81E-02** |
| **Plains** | 0.017 | 1.19E-01 |
| **Rocky Mountains** | 0.010 | 1.7E-01 |

**Supplementary Table 9**: Further Description/Lookup Dictionary of Covariates included in Linear Model

| Model Covariate Name | Description |
|---|---|
| Region 1 | New England |
| Region 2 | Mideast |
| Region 3 | Great Lakes |
| Region 4 | Plains |
| Region 5 | Southeast |
| Region 6 | Southwest |
| Region 7 | Rocky Mountains |
| Region 8 | Far West |
| Sex | 0=Female, 1=Male |
| Age=0 | 0 years |
| Age=1 | 1-4 years |
| Age=17 | 80-84 years |
| Age=18 | 85+ years |
| Hispanic | 0=Non-Hispanic, 1=Hispanic |
| Adjusted Race | 1=White, 2=Black, 4=Asian/Pacific Islander |

**Comparison of County Mortality Rates and Population Amongst Datasets**

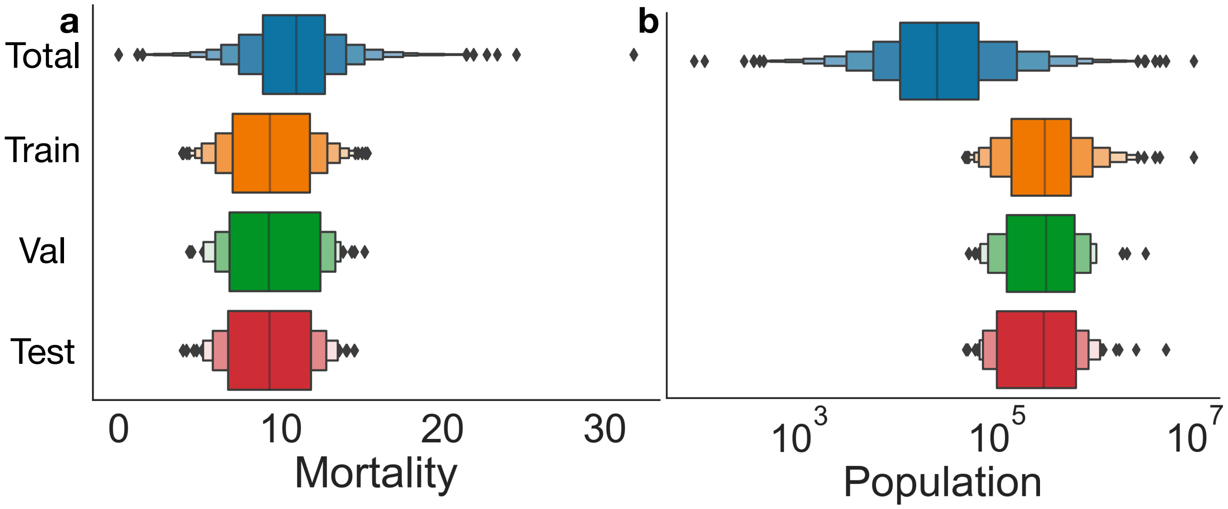


**Supplementary Figure 1:** Mortality and Population differences between all 3,142 total United States counties (Total), training (Train), validation (Val) and testing (Test) datasets visualized using boxenplots; a) Mortality; b) Population; while mortality is similar between all datasets, the 430 counties are far more populous than the Total set; selection criteria for the 430 counties featured in the study were designed to assign greater certainty in the mortality estimate (larger population) and sample a wider range of mortality estimates (inner boxes of boxenplots are wider than the Total); if sampling at random from Total, we would expect worse overall mortality estimates due to statistical imprecision and poor estimates for counties on the wide tails

#### Supplementary Overview of All Conducted Analyses

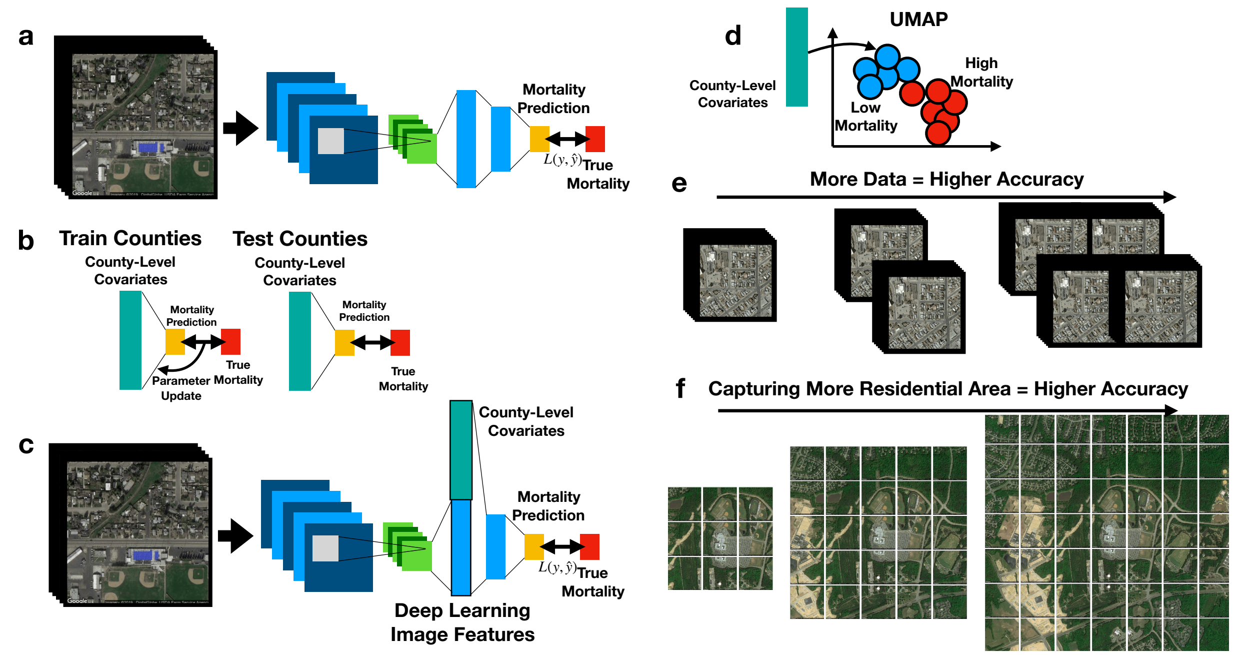


**Supplementary Figure 2: Graphical Overview of All Comparisons:** a) Image-only ResNet50 deep learning model, details on training, validation and testing featured in main text Figures 1-2; b) Linear regression model predicts mortality from test counties' county-level covariates after least-squares regression updating of parameters based on training dataset; c) combining county-level covariates with deep learning image features for prediction of mortality rate; training, validation and test workflow is identical to that of the image-only model, main text Figures 1-2, save for the addition of the county-level covariates to the deep learning features in the fully-connected output neural network layers; a complete overview of training, validation and testing can be found in Supplementary Figures 12-13; d) County-level covariates are used to describe clusters identified from Spectral Clustering UMAP embeddings from image-only model for test county images (Figure 2g); e-f) data scaling tests, see Supplementary Figure 11 for results; e) assessing the test set mortality prediction accuracy as a function of increasing the number of images/schools per county; f) assessing the test set mortality prediction accuracy as a function of incorporating more images from the surrounding neighborhoods around schools to focus more on the residential neighborhood and not the school

#### Further Description of Found SHAP and Convolutional Filter Features

Shadows under buildings, trees, and open highway space were associated with increased mortality, which could be an artifact of higher mortality rates in rural counties and possibly higher instances of foliage within these counties may produce more shadows. We found that the color contrast between the center and borders of roofs was associated with increased mortality. We hypothesize that lack of maintenance or longevity of the building can be attributed to aging or socioeconomic indicators which are also independently associated with mortality. Further inspection of learned image filters demonstrated the ability of our model to identify these factors by utilizing shape and light intensity.

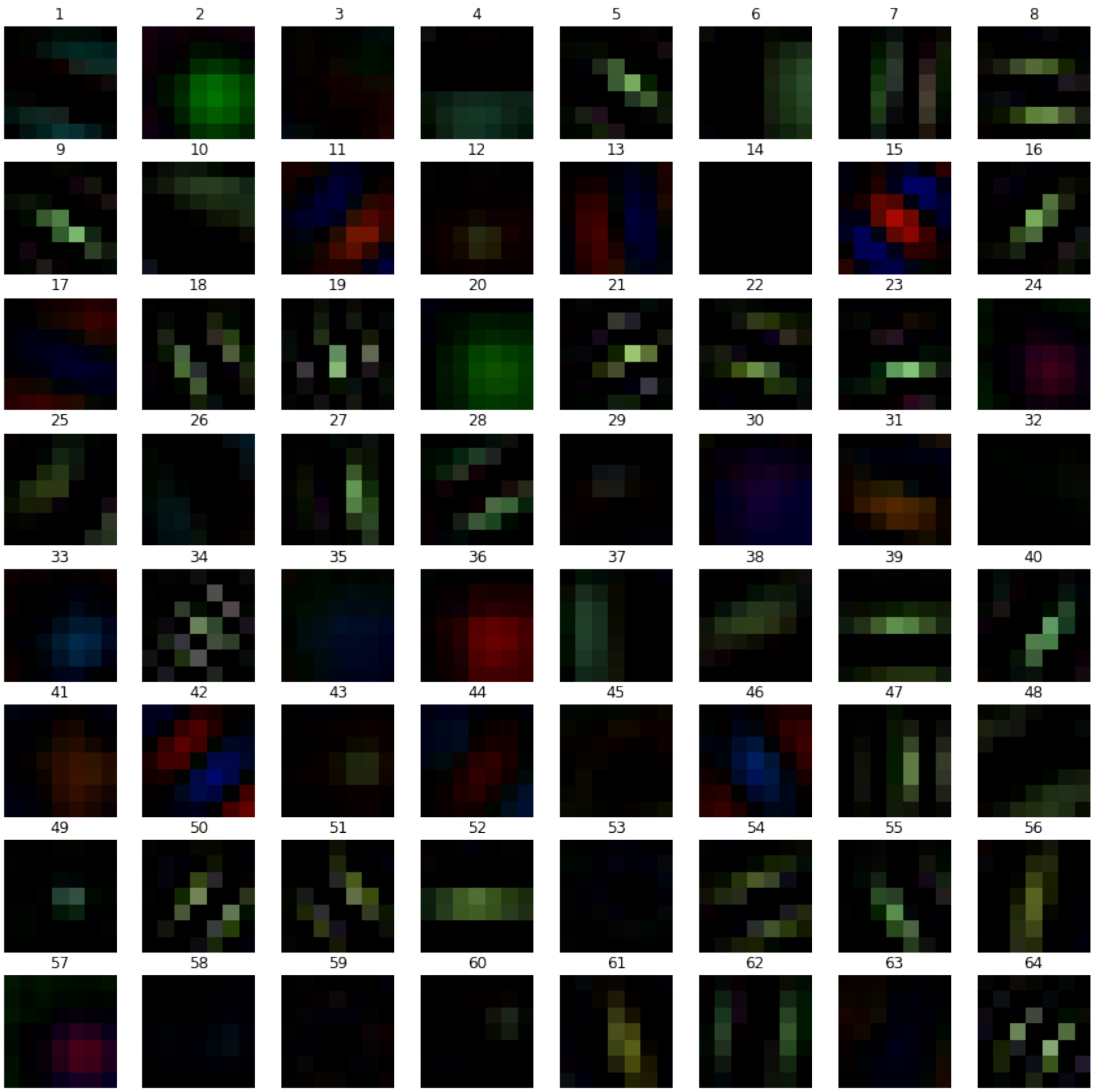


**Supplementary Figure 3:** Learned Convolutional Filters, numbered; we note the recurrence of patterns with parallel lines in different rotational orientations (filters 5, 8, 18, 22, 28, 50, 51, 54, 62, 64); similarities between filters 2, 6, 20; similarities between filters 15 and 42. Some filters place more weight on shape and morphology, while others focus more on color and intensity, as reflected in Supplementary Figures 4-6

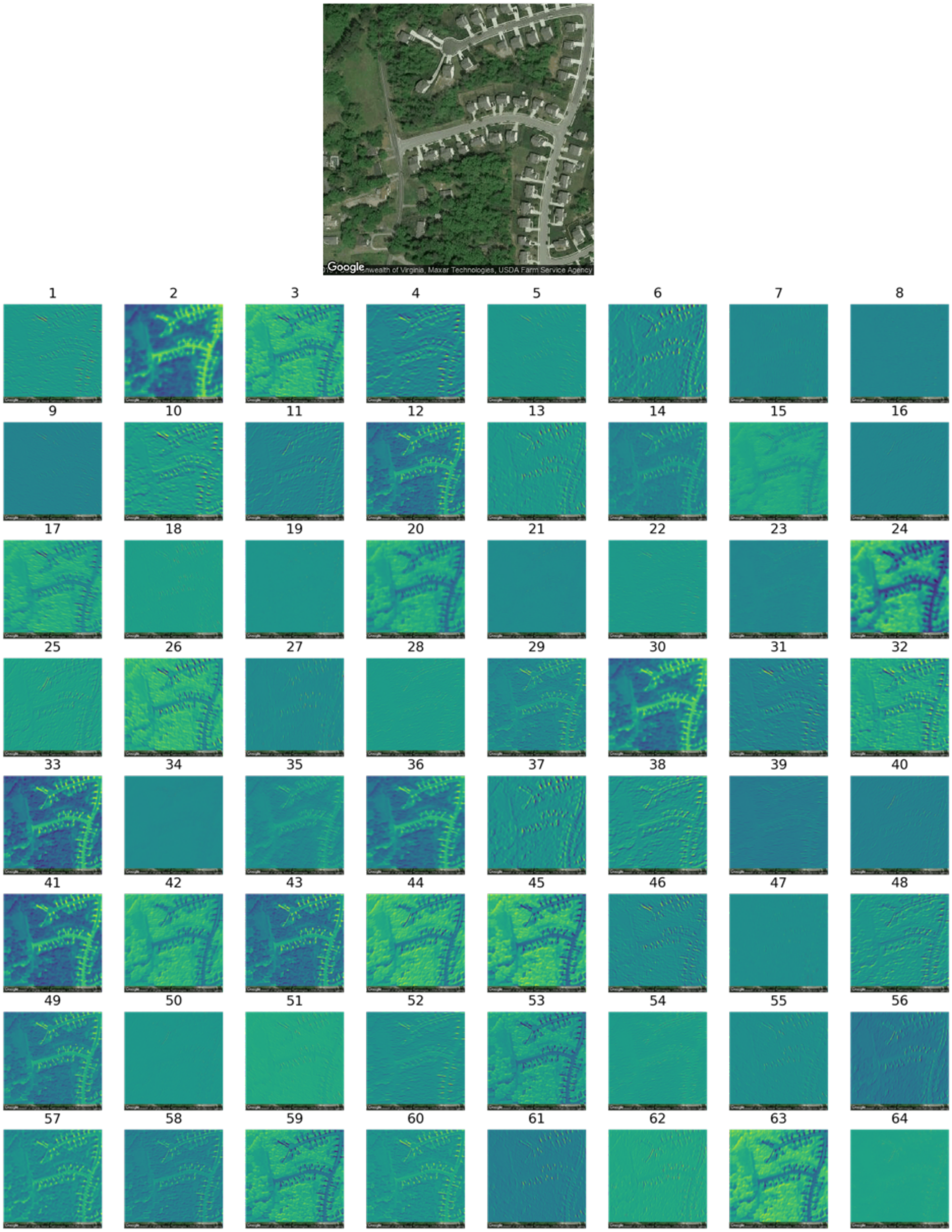


**Supplementary Figure 4**: Convolutional Filter Activations from VA41, Sampled Image around School 1; filter numbers correspond to the filters introduced in Supplementary Figure 3

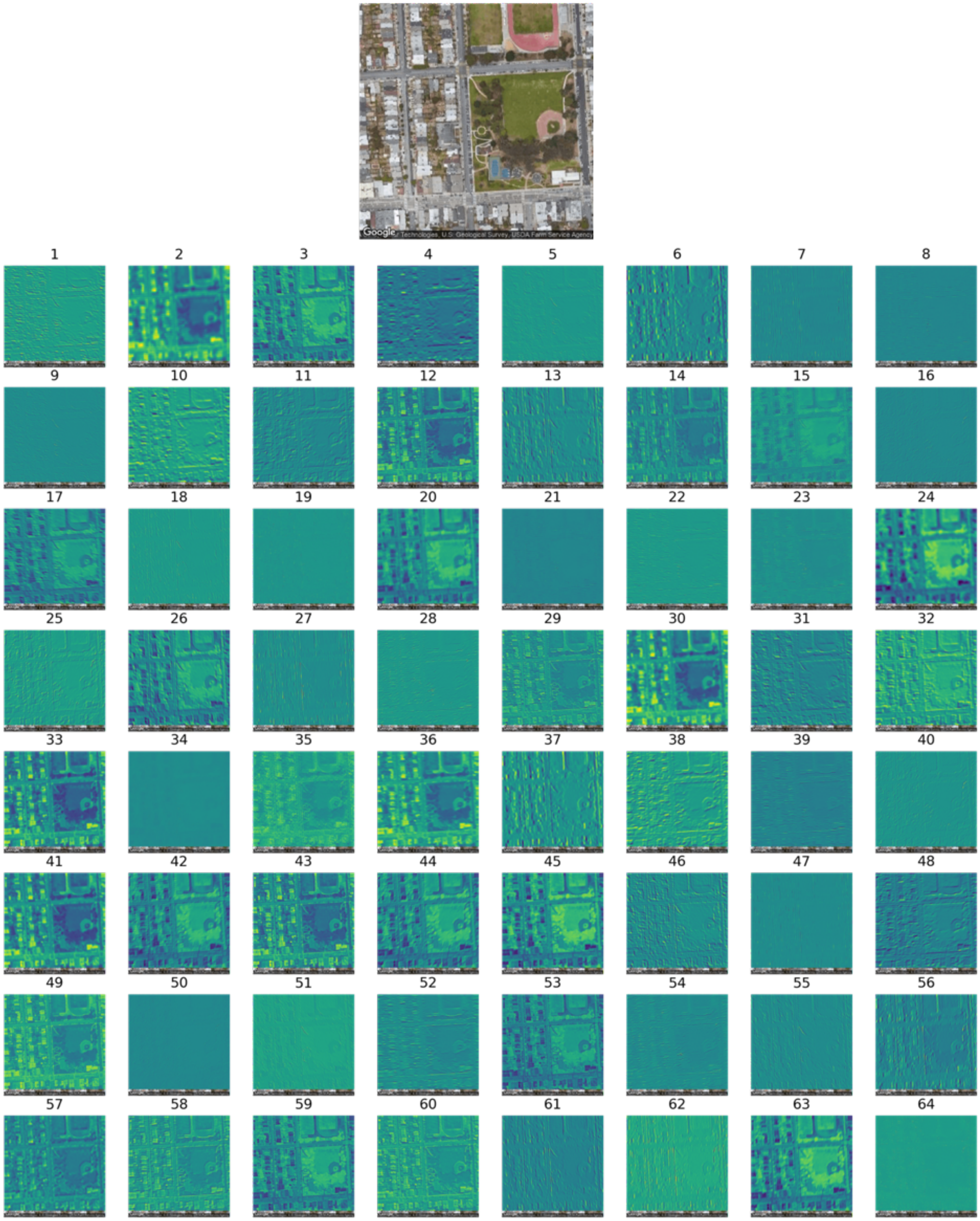


**Supplementary Figure 5:** Convolutional Filter Activations from CA75, Sampled Image around School 2; filter numbers correspond to the filters introduced in Supplementary Figure 3

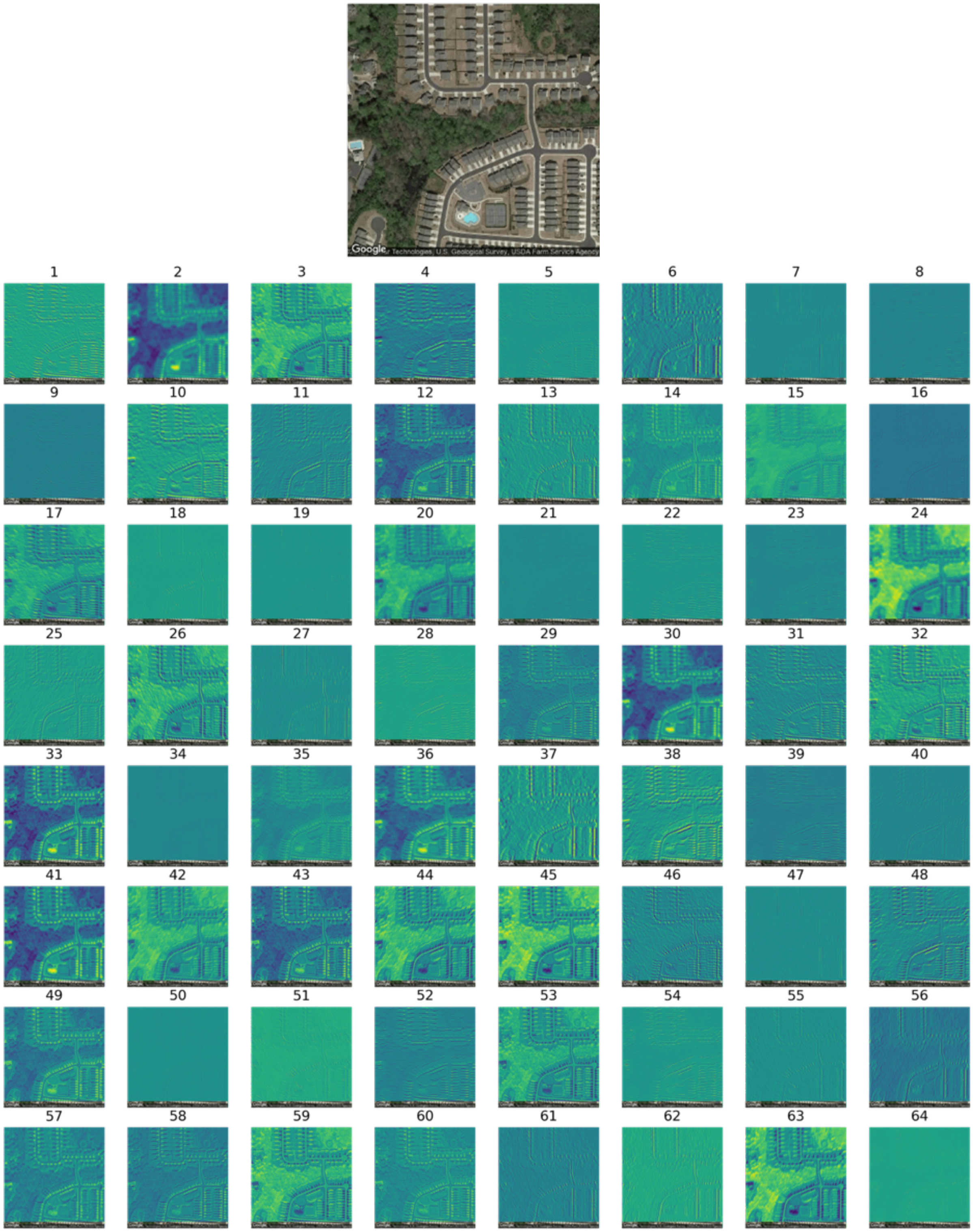


**Supplementary Figure 6**: Convolutional Filter Activations from CA75, Sampled Image around School 4; filter numbers correspond to the filters introduced in Supplementary Figure 3

**Further Description of Image Plot Embeddings**

We found that counties from certain regions tended to group together in the embedding plots, formed by 1000-d vectors extracted from each image using the neural network. We hypothesize that this is due to similar flora, landscaping practices, or similar distribution of schools throughout communities within the same region in the United States. The clustered image features demonstrated that satellite images across large geographic distances could also be related to other mortality predictors. For instance, we hypothesize that the images found in cluster 2 were associated with more affluent and well-educated suburban neighborhoods, given a high proportion of sidewalks and curved street design. Most of these images were taken from San Francisco County in California, where the average income and educational attainment were much greater than the averages found across the other clusters. One possible interpretation of the stark contrast in mortality rates between clusters 2 and 7 is that cluster 7 may have contained more images from a rural context. This is supported by the fact that the average county population significantly lower than all of the other clusters.

With regards to other mortality predictors, we noticed that cluster 5 image features, containing El Paso County, were associated with a large Hispanic population. Cluster 5 represented a county with moderate mortality, income and education, and primarily located in the southwestern and western United States. Of note here was a sub-cluster with a high Hispanic population, mostly comprised of images from El Paso County, Texas. The images within this sub-cluster were associated with lower mortality as compared to the rest of the image cluster. See supplementary figures 7-9.

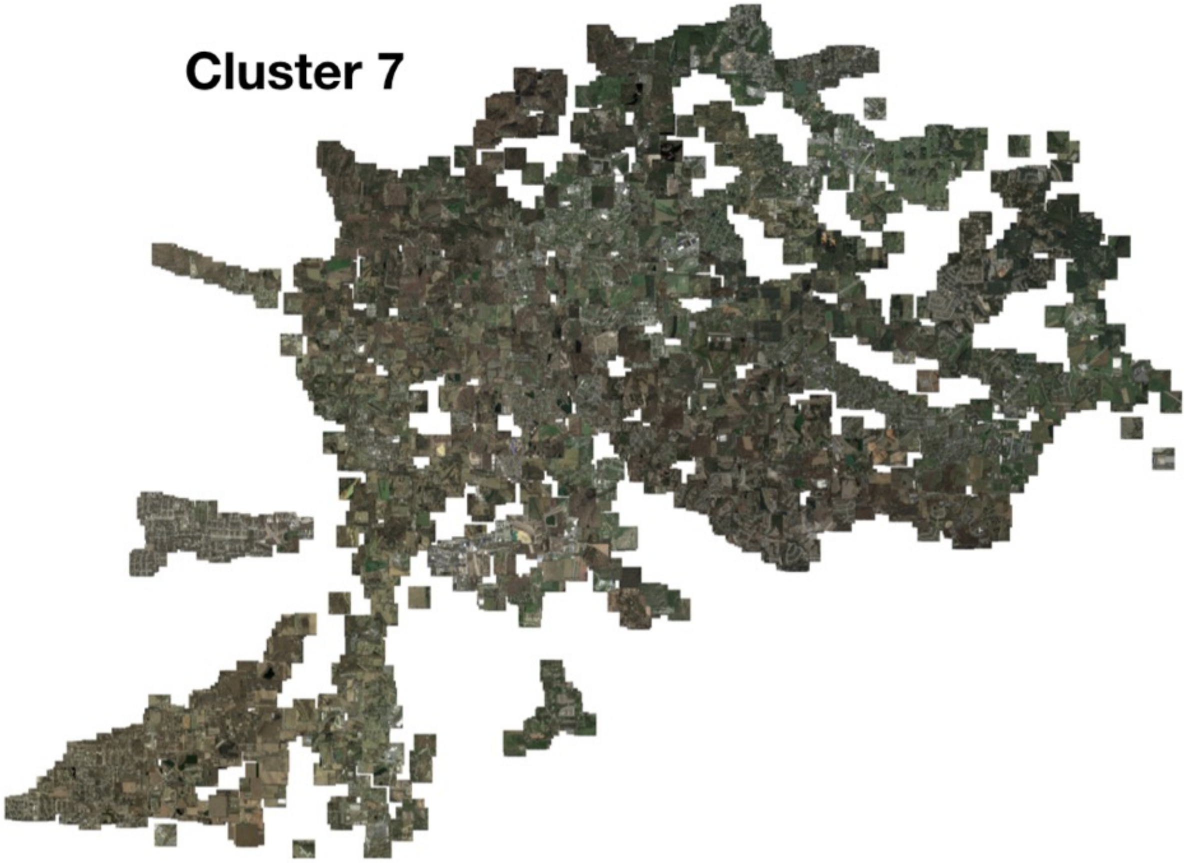


**Supplementary Figure 7:** UMAP Embeddings of Cluster 7 Overlaid with Original Images

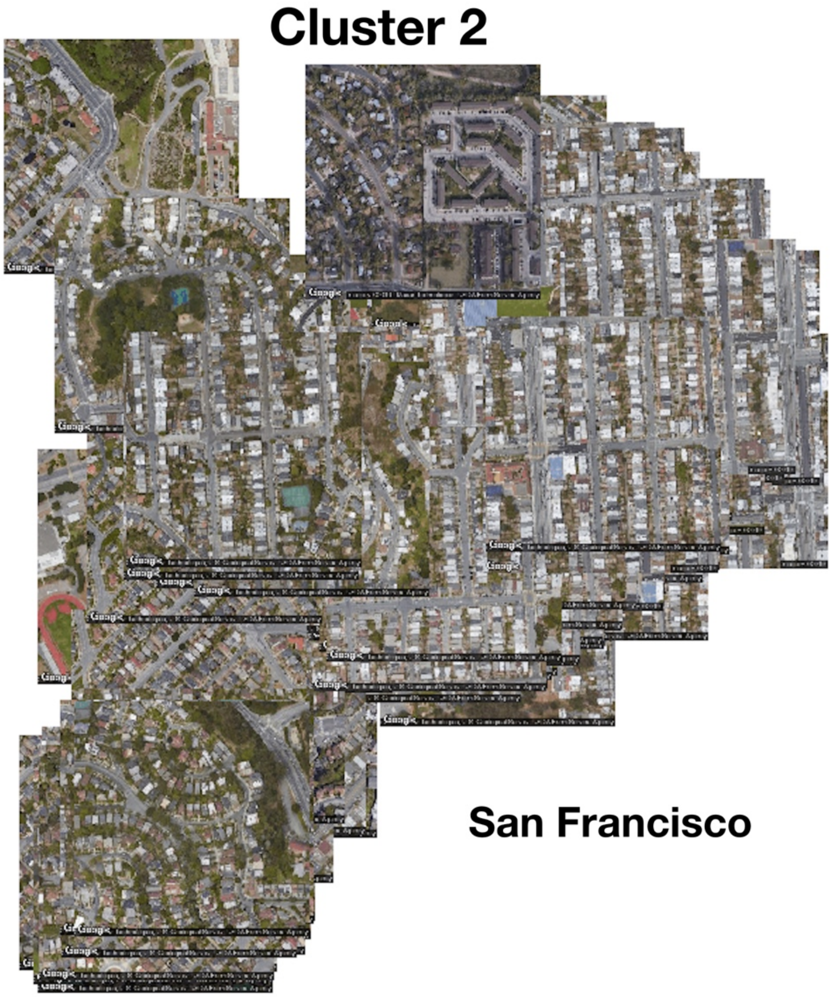


**Supplementary Figure 8:** UMAP Embeddings of Cluster 2 Overlaid with Original Images

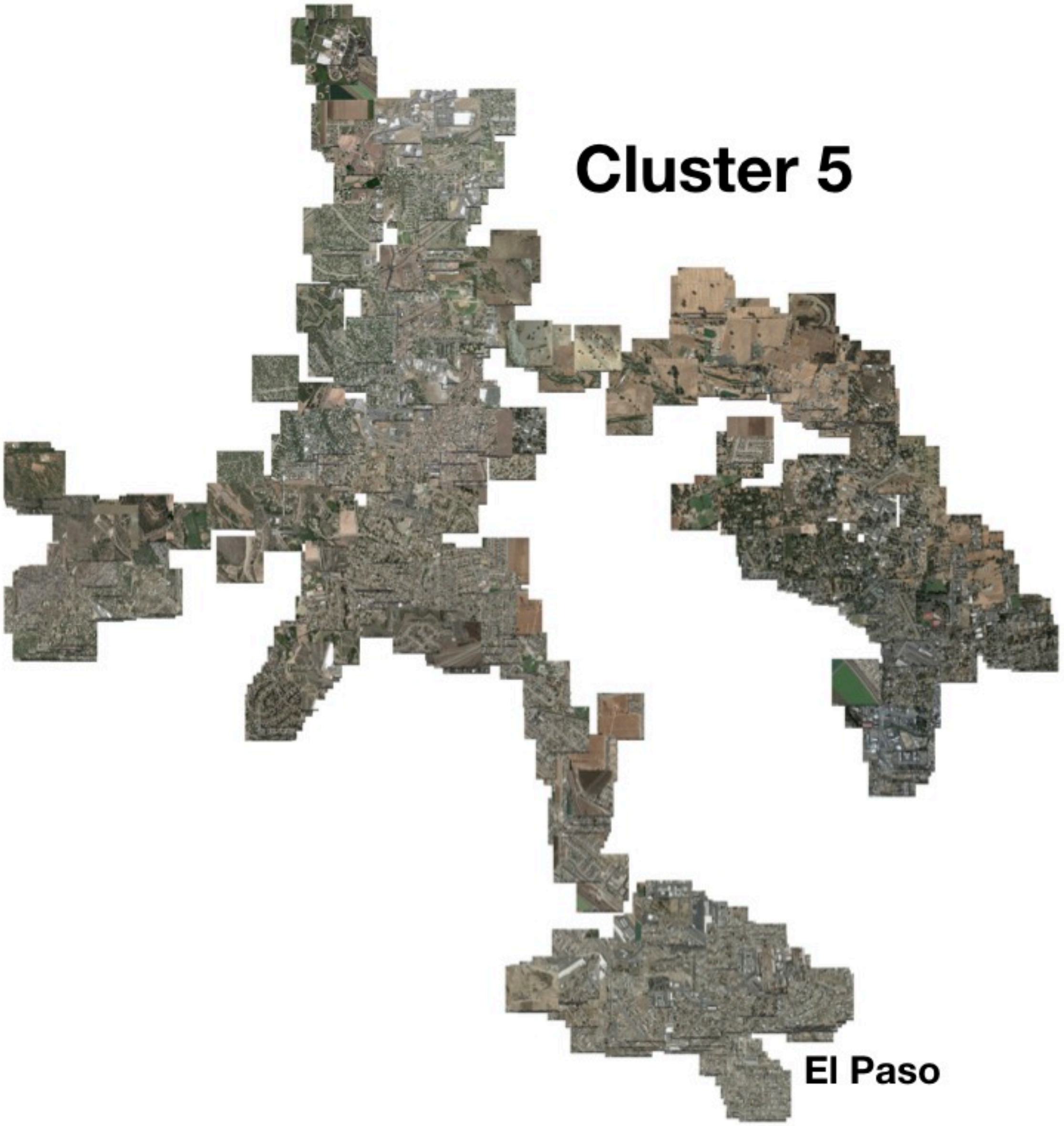


**Supplementary Figure 9**: UMAP Embeddings of Cluster 5 Overlaid with Original Images

#### Further Description of Image Clusters

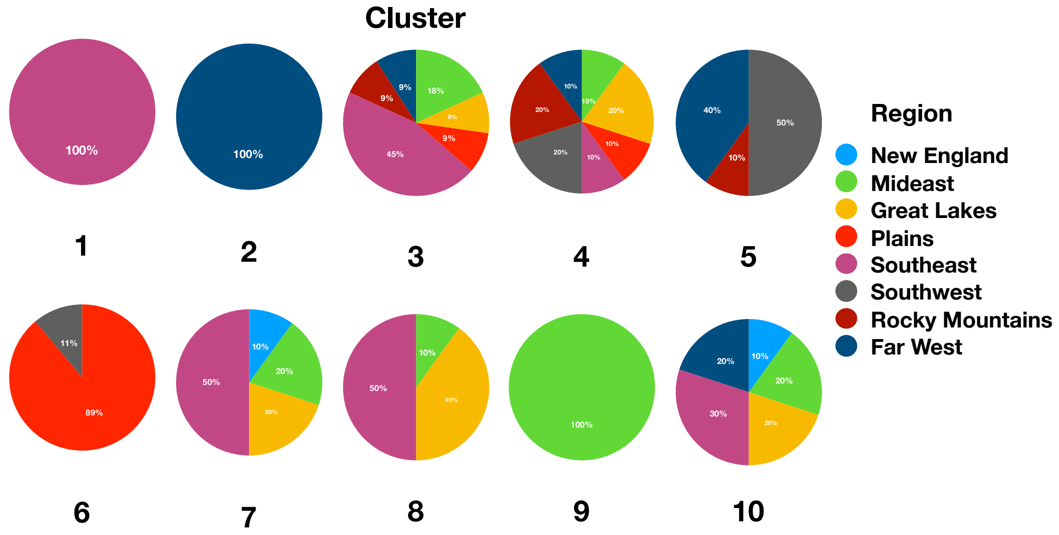


**Supplementary Figure 10:** Decomposition of Each Image Cluster by Region in US

**Effects of Sampling Larger Residential Areas Around Schools and Dataset Size on Predictions**

We assessed how sampling larger neighborhoods around schools and number of schools contributed to model predictive accuracy by varying both the sampling area (0-1 miles, 4 intermediate values depending on number of images) and number of schools (1-4) simultaneously using a grid search and reporting the test set pearson correlation between predicted and true county level mortality rates. We estimated the relationship between sampling area, number of schools as the independent variables and the pearson correlation as the dependent variable, modeled the interaction between the sampling area as a monotonic ordinal predictor and number of schools as a categorical predictor to derive conditional effects using the Bayesian modeling package *brms* [1,2]. Results indicate that when fixing the number of sampled schools to 2 or 3 ($\beta_2 = 0.039[95\%CI: 0.028,0.048]$; $\beta_3 = 0.023[95\%CI: 0.023,0.042]$), the predictive correlation sharply increases with larger area sampled, yet this does not occur when the number of schools are 1 or 4 ($\beta_1 = -0.002[95\%CI: -0.010,0.007]$; $\beta_4 = 1.237e-4[95\%CI: -0.009,0.010]$), which could indicate insufficient data in the former case and saturation of the effect as a function of number of images in the latter case. Note that we did not subsample images during this grid search to control sample size, so we cannot rule out that the increases in predictive accuracy were driven by increases in number of training images. Number of images per county are a function of the sampling area per school and number of schools sampled per county.

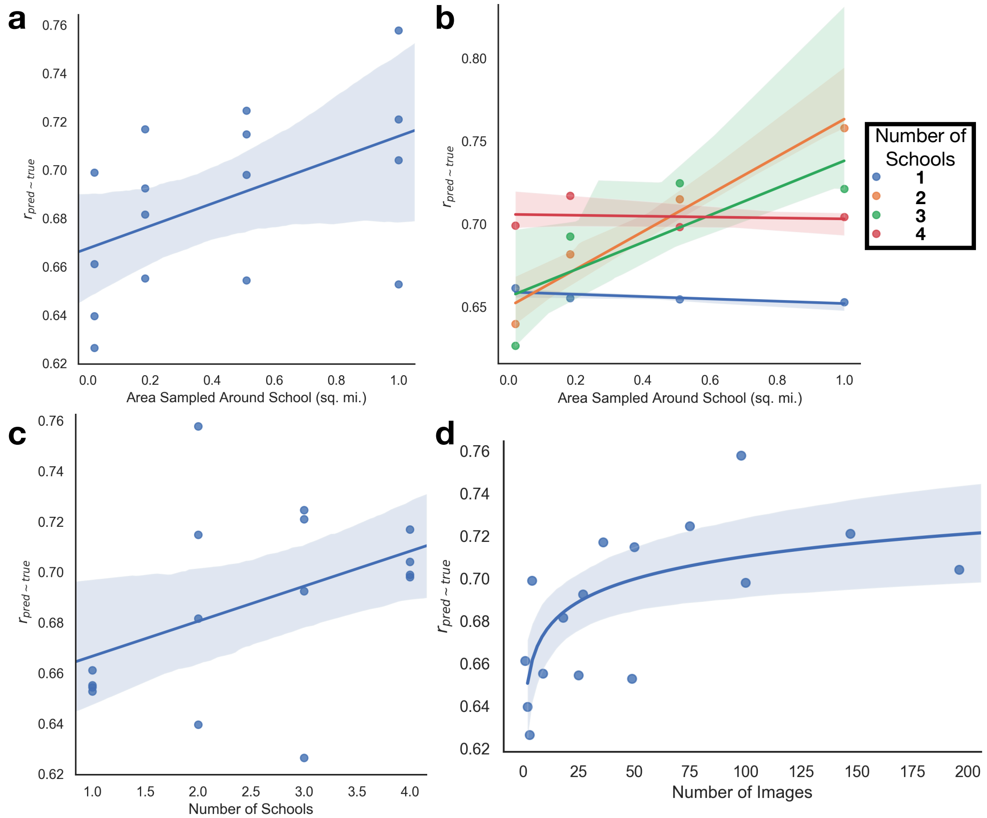


**Supplementary Figure 11**: **Dataset Scaling Tests: (a)** Regression plot of area sampled around each school versus the correlation between predicted and true mortality of the validation counties; **(b)** Regression plot of area sampled around each school versus the correlation between predicted and true mortality of the validation counties, colored by number of schools; **(c)** Regression plot of number of schools selected per county versus the correlation between predicted and true mortality of the validation counties; **(d)** Regression plot of the number of images sampled for each county versus the correlation between predicted and true mortality of the validation counties; $r_{pred \sim true}$ represents the pearson's correlation coefficient between predicted and true mortality of the validation counties

#### Covariate Adjustment During Deep Learning Model Training and Evaluation

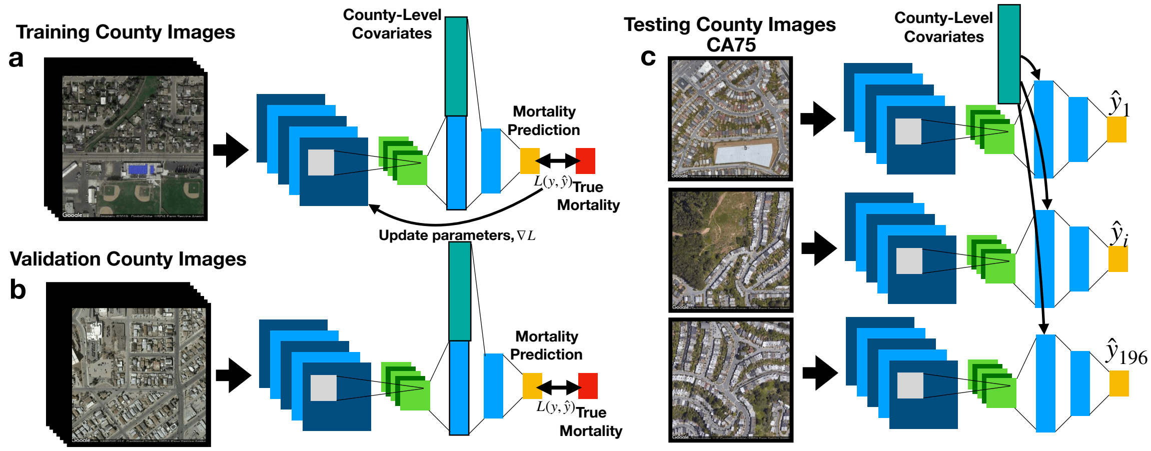


**Supplementary Figure 12: Overview of Deep Learning approach that combines image and county-level covariate features, with workflow identical to main text Figure 2:** a) training images pass through pre-trained (Figure 2b) ResNet50; county-level covariate information represented as vector combined with image features using a Kronecker's Outer Product in the output fully connected layers to predict county-level mortality, compared to true mortality to update model parameters; d) during training, ResNet50 model predicts mortality from combined image and county-level covariate data from validation counties to estimate validation loss, which is compared to training loss over training epochs to determine when training should stop to avoid overfitting; e) trained ResNet50 model predicts county-level mortality on 196 individual images (one mortality rate prediction per image; county-level covariate information separately combined with each of the individual images using same operations on training/validation sets) from each test county; for each test county, 196 image-level mortality rate predictions are averaged using a trimmed mean to infer the overall county-level mortality rate; the overall county-level mortality rate is correlated to true county-level mortality rate

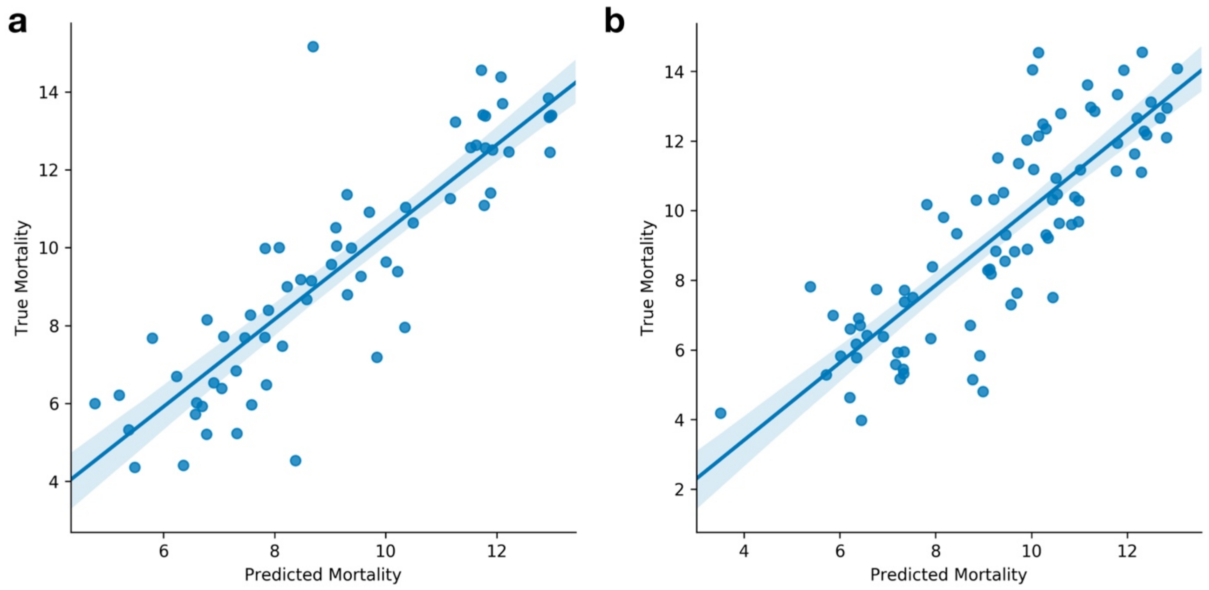


**Supplementary Figure 13**: **Preliminary Tests with Covariate Adjustment, Predicted vs True Mortality for: (a)** Validation Counties (n=65), **(b)** Test Counties (n=86) ; Deep Learning Covariate-Adjusted Model predicts mortality on the image level, each image's embedding is

adjusted with county level demographic information, interactions were modeled between the image features and demographic information with a Kronecker's Outer Product; Predictions are derived for each image and averaged across the county images to yield a final prediction for county level mortality
